# Physical Activity Behaviour in Middle-Aged and Older Canadian Women and Men: An Analysis of the CLSA

**DOI:** 10.1101/2025.03.17.25323990

**Authors:** C D’Amore, LE Griffith, J Richardson, MK Beauchamp

**Author notes:** **Corresponding Author:** Dr. Marla Beauchamp, Associate Professor, School of Rehabilitation Science, Canada Research Chair in Mobility, Aging and Chronic Disease, McMaster University, 1400 Main Street West, Hamilton, Ontario, Canada, L8S 1C7.

## Abstract

**Background:** Increasing physical activity (PA) levels can decrease the burden of non-communicable diseases and improve functional ability in aging populations. Understanding current patterns in PA behaviours is essential for developing effective interventions. This study aimed to describe the usual PA by type of activity and amount in middle-aged and older Canadians.

**Methods:** A descriptive cross-sectional analysis of baseline data from the Physical Activity Scale for the Elderly in the Canadian Longitudinal Study on Aging was completed. Subgroup analyses were used to explore PA behaviour, by age and sex, socioeconomic variables, region and season. In addition, we estimated quintiles based on the amount of total PA completed in Canadians 45-85 years. Means and frequencies were reported using inflation weights.

**Results:** The 47,840 participants represented our target population of 12,365,513 Canadians 45-85 years old. The mean PASE score was 151 (SD 79.11) with 65% of Canadians 45-85 years completing at least 150-minutes of moderate-vigorous PA a week. Amount of PA and the proportion of Canadians meeting the recommendation decreased for females, and with increasing age, lower income, and less education. Additionally, those with the lowest PA levels were more likely to report limitations in mobility and activities of daily living and had a higher prevalence of some chronic conditions (diabetes, musculoskeletal, and vision).

**Conclusion:** Physical activity behaviour among middle-aged and older Canadians varies based on several characteristics. Targeted interventions and promotion efforts are warranted, particularly for older females and those with lower income and education. Further investigation to determine directionality is needed.

## INTRODUCTION

Physical activity is an important modifiable behaviour for healthy aging.^1^ Defined as any bodily movement produced by skeletal muscle that requires energy expenditure,^2^ physical activity (PA) levels vary across countries, age groups and sex, but it is estimated that over a quarter of adults worldwide are not achieving the recommended levels of PA for optimum health (150-minutes of moderate-to-vigorous PA).^3^ Increasing PA levels has the potential to decrease the economic burden on healthcare systems by over $500 million per year in Canada and $37 billion CAD worldwide.^4, 5^ Understanding the patterns of PA behaviours and identifying individuals at greater risk for low levels of PA is essential for informing interventions and health promotion efforts.

Physical activity literature has begun to shift in focus from higher-intensity activities, referred to as moderate-vigorous (MVPA), to including the full spectrum of movement behaviours in daily life. The importance of lighter intensity activities (i.e., activities that do not elevate heart rate or cause perspiration)^6^ is increasingly recognized in the discourse on PA and health and mortality,^7, 8^ and national guidelines like the Canadian 24-hour movement guidelines.^9^ Promoting a wider range of intensities as part of PA promotion is particularly relevant for populations who favour lighter intensities (e.g., older adults and women),^10^ allowing for a more inclusive consideration of PA behaviour patterns that contribute to health. Therefore, it is extremely important to use measurement tools that capture a broad spectrum of physical activities.

Although low PA levels are considered a global health crisis, higher-income countries like Canada are said to experience a greater relative burden of inactivity.^11^ Nonetheless, there is a dearth of studies that have comprehensively examined PA levels (i.e., include all intensities rather than only MVPA) among Canadian adults. This work describes the spectrum of usual PA (i.e., light to vigorous intensities) by type of activity and amount in middle-aged and older Canadians. Specific aims were to: i) describe PA behaviour in Canadians aged 45-85 years, including by important subgroups related to age,^12^ sex,^12^ socioeconomic status,^13^ and region and season;^14^ and ii) describe Canadians by the amount PA undertaken. Identifying subpopulations at greater risk of having lower PA, and patterns of PA behaviour, can inform targeted health promotion efforts for aging populations in Canada and similar high-income countries.

## METHODS

### Study Population

The Canadian Longitudinal Study on Aging (CLSA) is a population-based study of 51,338 Canadians across all 10 provinces. Baseline data collection was completed in 2015; participants were 45-85 years old and free from any cognitive impairment at the time of recruitment.^15, 16^ More details on sampling and exclusion criteria of the CLSA are available in appendix A. As part of the baseline data all participants were contacted approximately 1.5 years after their initial assessments to complete a maintaining contact questionnaire (MCQ), to help improve participant retention and administer several additional modules including PA.^16^ A total of 47,840 completed the MCQ; 3,498 participants did not complete this additional baseline assessment and were not included in the present analysis.

### Physical Activity

The Physical Activity Scale for the Elderly (PASE) is a 7-day retrospective questionnaire designed to be more representative of activities undertaken by older people.^17^ Participants report on walking, light sport and recreation, moderate sport and recreation, strenuous sport and recreation, exercise, light housework, (indoor and outdoor), heavy housework (indoor and outdoor), home repairs, caregiving roles, work or volunteer activities, and sedentary. Details on the activity types and questions can be found in the supplemental table B2. The CLSA administered a slightly modified version of the PASE as part of the MCQ.^18^ Question responses to the PASE were used individually and together to operationalize four PA outcomes.

#### PASE total score

was calculated for all participants with the required information available, rounded to a whole number as per the PASE instruction manual.^19^

#### Proportion of total score

The average proportion of total score for each activity was reported. Values range from 0 (do not contribute to total PA score) to 1 (is the sole contributor to total PA score). To determine the proportion of total PA score represented by each activity, each activity score (calculated from weights and question responses) was divided by the total score for each person.

#### Prevalence of activity types

The percentage of participants who completed each activity in the previous 7-days was calculated.

#### 150-minutes of MVPA guidelines

The total time spent in walking, moderate recreation, strenuous recreation, and exercise were calculated using the methods by Mayo et al. (2021), details available in supplemental appendix B.^20^ Individuals with at least 150-minutes of MVPA in the last 7-days were said to have met the guideline.

### Stratification

In total, six sets of subgroups based on correlates of PA were explored for aim 1: i) age and sex (eight subgroups), ii) household income (five subgroups), iii) education (four subgroups), iv) material deprivation (5 subgroups), v) social deprivation domain (5 subgroups) and vi) region and season (16 subgroups; four regions and four seasons). Subgroup details can be found in appendix C. For the second aim, participants were grouped into weighted quintiles based on total PASE score to describe amount of PA, where Q1 represents those with lowest 20% of PA levels and Q5 representing those with the highest 20% of PA levels.

### Statistical Analysis

Analyses were completed using R studio (Rversion 4.1.2). Inflation sampling weights provided by the CLSA were used to estimate descriptors for the target population (R package ‘*Svy*’). ^21^ Sample and population descriptors were calculated using mean (standard deviation (SD)) and frequency (percent). Weighted and unweighted estimates of demographic, health, clinical, social, and environmental characteristics were calculated for the analytic sample. Physical activity behaviour was described using the four PA outcomes for each of the designated subgroups in aim 1. For aim 2, proportions and means for each PA quintile were estimated for correlates (e.g., age, sex, chronic conditions, environment). Missing data and patterns of missingness were investigated (R package ‘*mice*’). Pairwise deletion was used, meaning the sample size varied between analyses based on which variables were included. Research ethics approval for this secondary analysis was obtained from the Hamilton Integrated Research Ethics Board (application #7342).

## RESULTS

### Sample and Target Population

The 47,840 participants represented a population of 12,365,513 Canadians 45-85 years old. Six-hundred and thirty-three participants (1.3%) were missing information required to calculate the total PASE score. Our estimated population was 51.8% female, and 95.0% reported being of European descent. Over 75.0% of the target population was taking at least one prescription medication, more than 85.0% reported good or better health, over half received a degree or diploma, and 75.2% were in a relationship. The average PASE score was 151 (SD79.11) with 64.8% of Canadians 45-85 years meeting the 150-minutes of MVPA. Most participants (∼70%) reported that the PA captured over the previous 7-days on the PASE was representative of their PA over the last 12 months. Full details on the sample, target population, and missing data are available in the appendix D.

### General Patterns in Physical Activity Behaviour

#### Age and Sex

Total PASE scores decreased with increasing age and were consistently lower in females (approximately 20-points lower in each age group). Figure 1 presents the proportion of total PASE score from each activity; for example, work made up a greater proportion of overall PASE scores for younger ages. The proportion of the score made up by light indoor housework increased with age, and heavy outdoor activities appeared to account for a larger proportion of scores in males than females. Exploring the percent of people who participated in each activity revealed similar results; home repairs and heavy outdoor activities were more common for males compared to caregiving which appeared to be more common for females. The youngest age group had the highest percent of people participating in strenuous recreation activities. Walking and light indoor housework were the most common activities across age groups and sexes.

**Figure 1.**
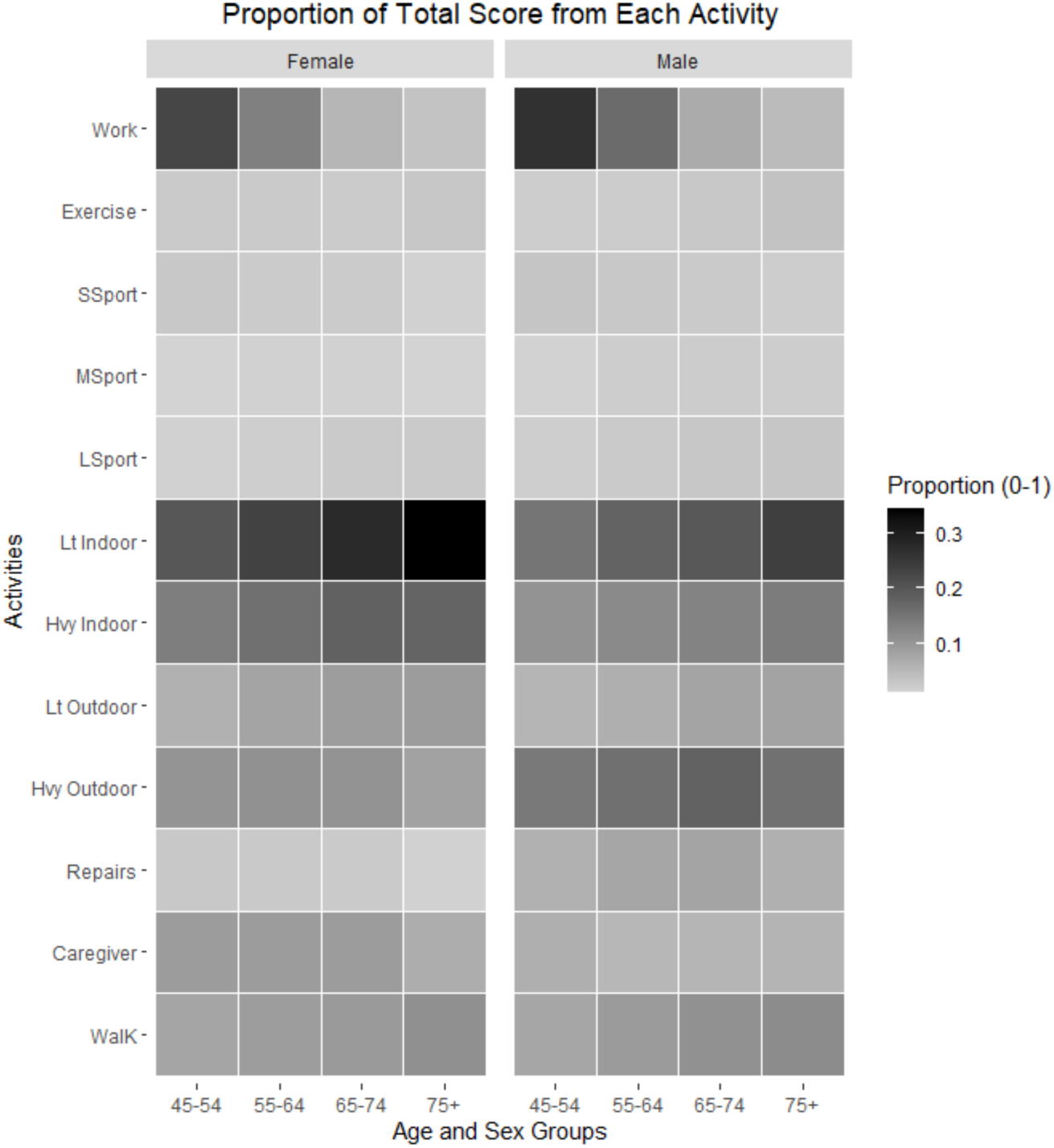
Proportion of PASE Scores for Males and Females. Each square represents the average proportion of the total PASE contributed by that activity for the subgroup. Where darker squares represent a greater proportion of the total score for that activity.

Additionally, the percent of people reaching 150-minutes of MVPA decreased in both males and females with age (details available in appendix E).

#### Household Income

Total PASE score increased with household income; however, the increase was most prominent between the two lowest income groups (28-point difference). When accounting for age, the increases in PASE score between income groups was smaller in higher age groups. For example, the 45-54 year old age group had a 44-point difference between the <$20,000 and the $20,000-$50,000 groups compared to the 17-point difference for the 75+ age group between the same income categories (Figure 2).

**Figure 2.**
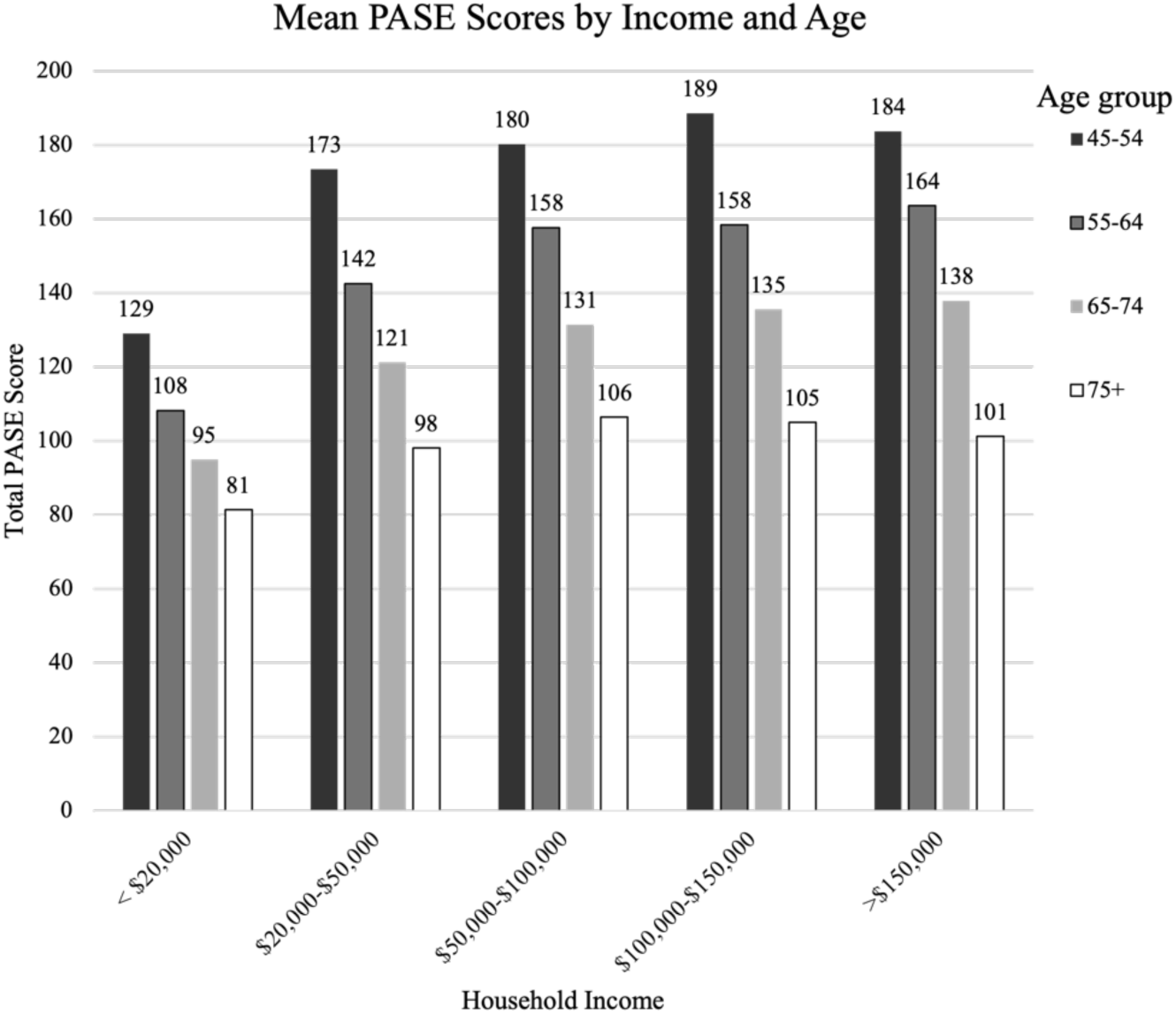
Mean PASE Scores Across Age Groups for Each Household Income Level.

Work appeared to make up a larger proportion of PASE scores for those with higher incomes; whereas, the proportion of total score from indoor housework decreased with higher income. However, after accounting for age, these patterns were less prominent across income groups and more pronounced when looking at age groups (Figure 3). There was only one noticeable difference for the prevalence of each activity, a higher percentage of individuals in the ≥$100,000 income group participated in strenuous recreation activities. An increase in the percentage of Canadians reaching 150-minutes of MVPA with increasing income level was consistent even after accounting for age. More details available in appendix F.

**Figure 3.**
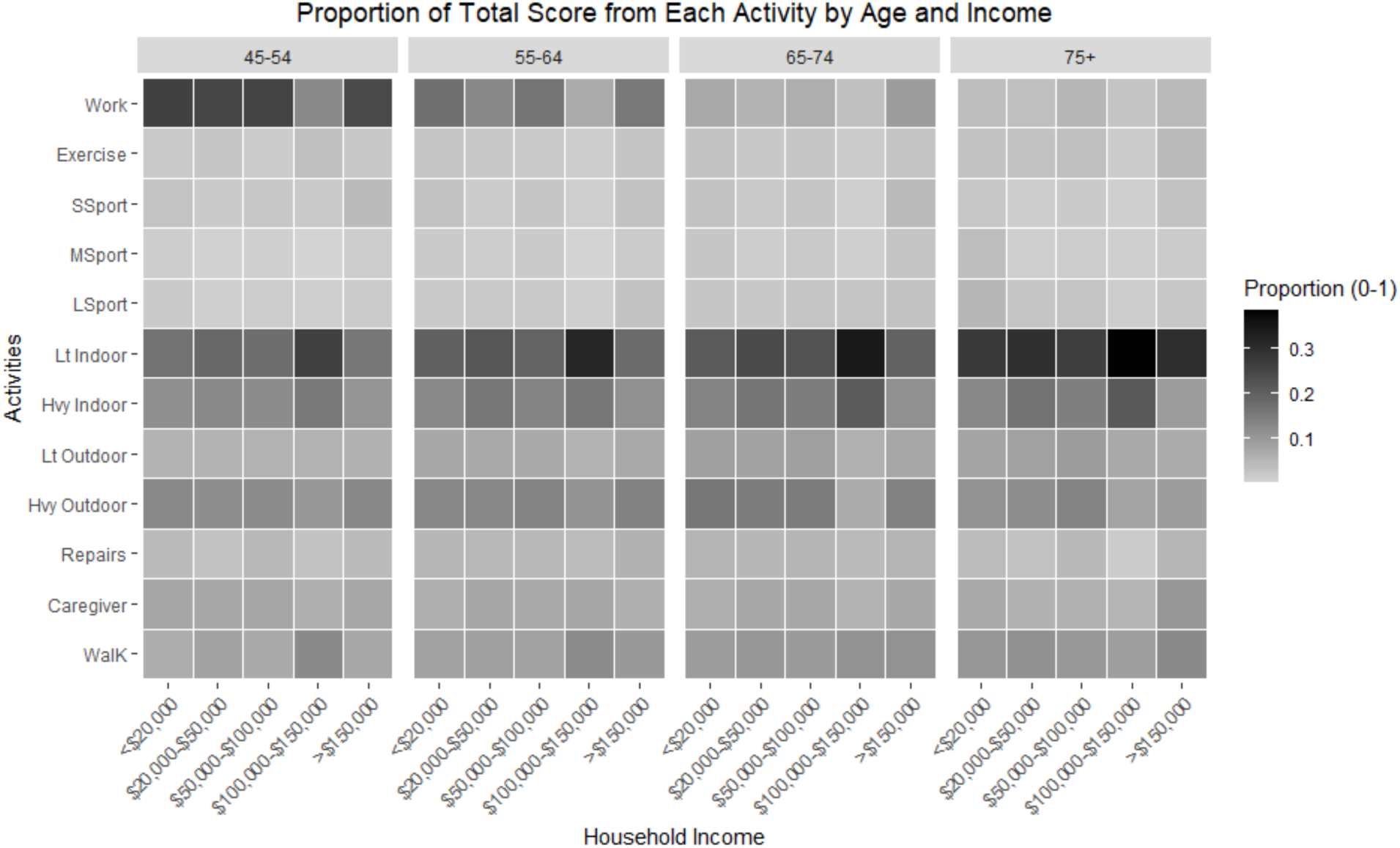
Proportion of PASE Score by Age Groups Across Household Income. Each square represents the average proportion of the total PASE contributed by that activity for a subgroup. Darker squares represent a greater proportion of the total score for that activity.

#### Education

Total PASE scores increased with education level. The largest increase was seen between those who have graduated and have not graduated from secondary school (17-points). Only minor differences were seen for proportion of score and percent of people completing each activity, with no clear overarching patterns (appendix G). As with total score, the percent of Canadians who achieved recommended levels of MVPA increased with education level; the largest jump was between the two lowest education groups (9.0%).

#### Material and Social Deprivation Index

Material deprivation scores had less of a pattern in total PASE score with the highest mean score in Q2 (157 SD81.37) and the lowest score in the most deprived group (Q5; 146 SD79.39). A clearer pattern was seen for social deprivation score where PA levels decreased with increasing deprivation. No strong patterns were seen for PASE score proportions or types of activities completed for either deprivation scale. The percent of people reaching 150-minutes of MVPA was similar across quintiles for both domain (difference between Q1 and Q5 material 7%, social 2%, appendix H).

#### Region and Season

Mean total PASE score varied between regions with residents in the Prairies (156 SD81.69) having the highest followed by British Columbia (152 SD78.82), Central (150 SD78.45) and Atlantic Canada (147 SD79.19) having the lowest. However, when season was considered, the Prairies only retained the highest PA amount for Jan-Mar, with Central Canada having the highest from Jul-Sep, and British Columbia having slightly higher PASE scores for the remaining two seasons (Figure 4). Atlantic Canada remained the lowest PA levels three of four seasons with the most pronounced differences in the colder months.

**Figure 4.**
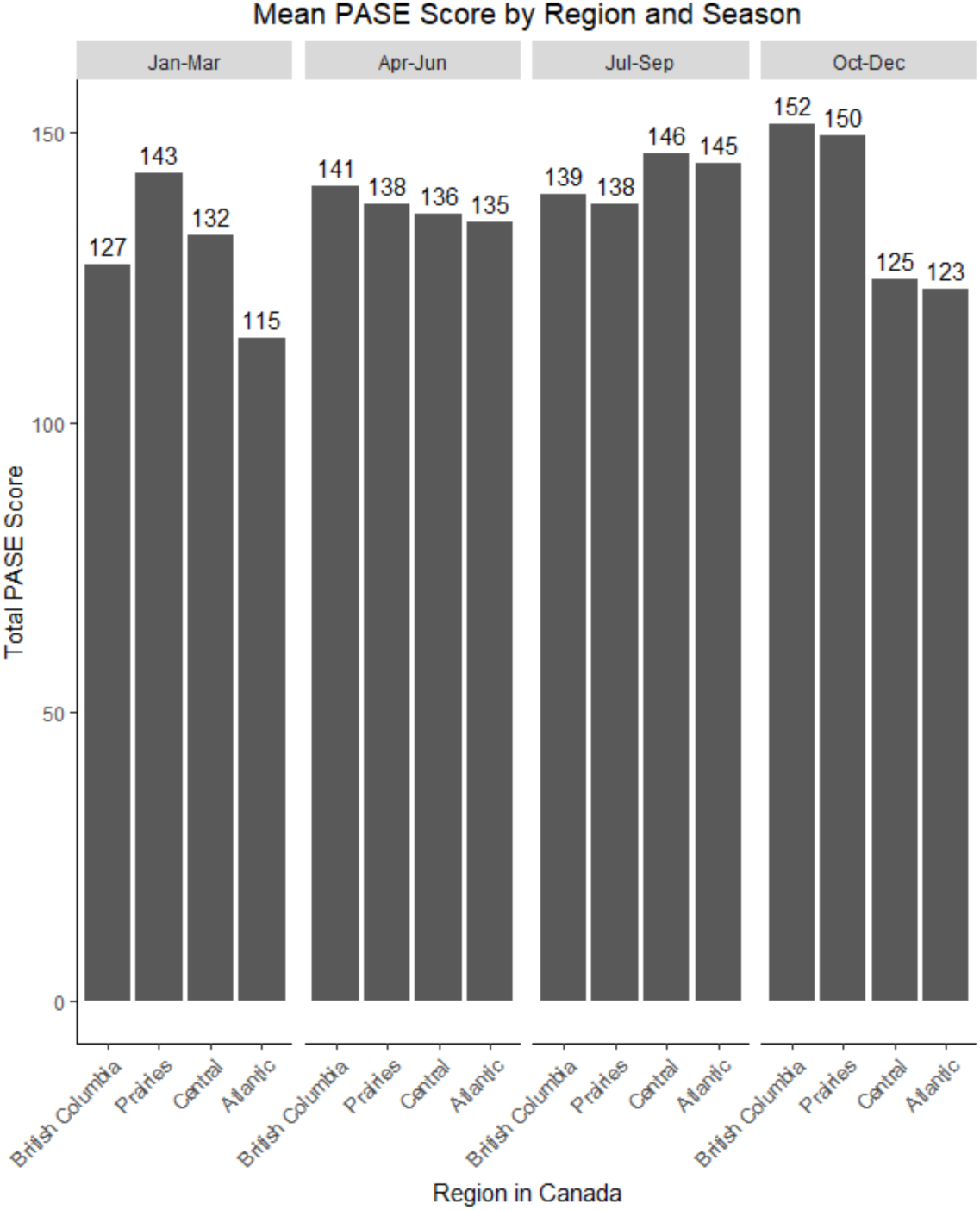
Variations in mean PASE scores between regions and seasons in Canada.

The Prairies had a higher proportion of their PASE total score coming from work and British Colombia had a smaller proportion of total score from heavy outdoor housework. Regions were very similar in the percentage of people completing each type of activity. British Columbia has a slightly higher percentage of people completing 150-minutes of MVPA a week (71%) compared to the other three regions (61-64%). Visual depictions are available in appendix I.

### Physical Activity Behaviour

#### Physical Activity Level Quintiles

Physical activity quintiles were: Q1 ≤85, Q2 86-121, Q3 122-158, Q4 159-217, and Q5 ≥218 (Table 1). The percent of females gradually decreased with increasing PA quintile. There was a 10% difference in the percent of people making over $150,000 per household between the lowest and the highest quintile. There was a 15.0% shift from those with no secondary school graduation to those with a degree or diploma when going from Q1 to Q5. The prevalence of diabetes, musculoskeletal, neurological and vision conditions was at least 10% higher in the least active compared to the most active. Almost a quarter (22.7%) of individuals in Q1 reported fair or poor general health, which was reduced to 12.9% in Q2 and only 7.5% in the most active group. There was over a 15.0% decrease in the presence of an ADL/IADL limitation going from Q1 to Q5. Disparities of PA across household income, education, self-reported health, and sex can be seen in Figure 5.

**Figure 5.**
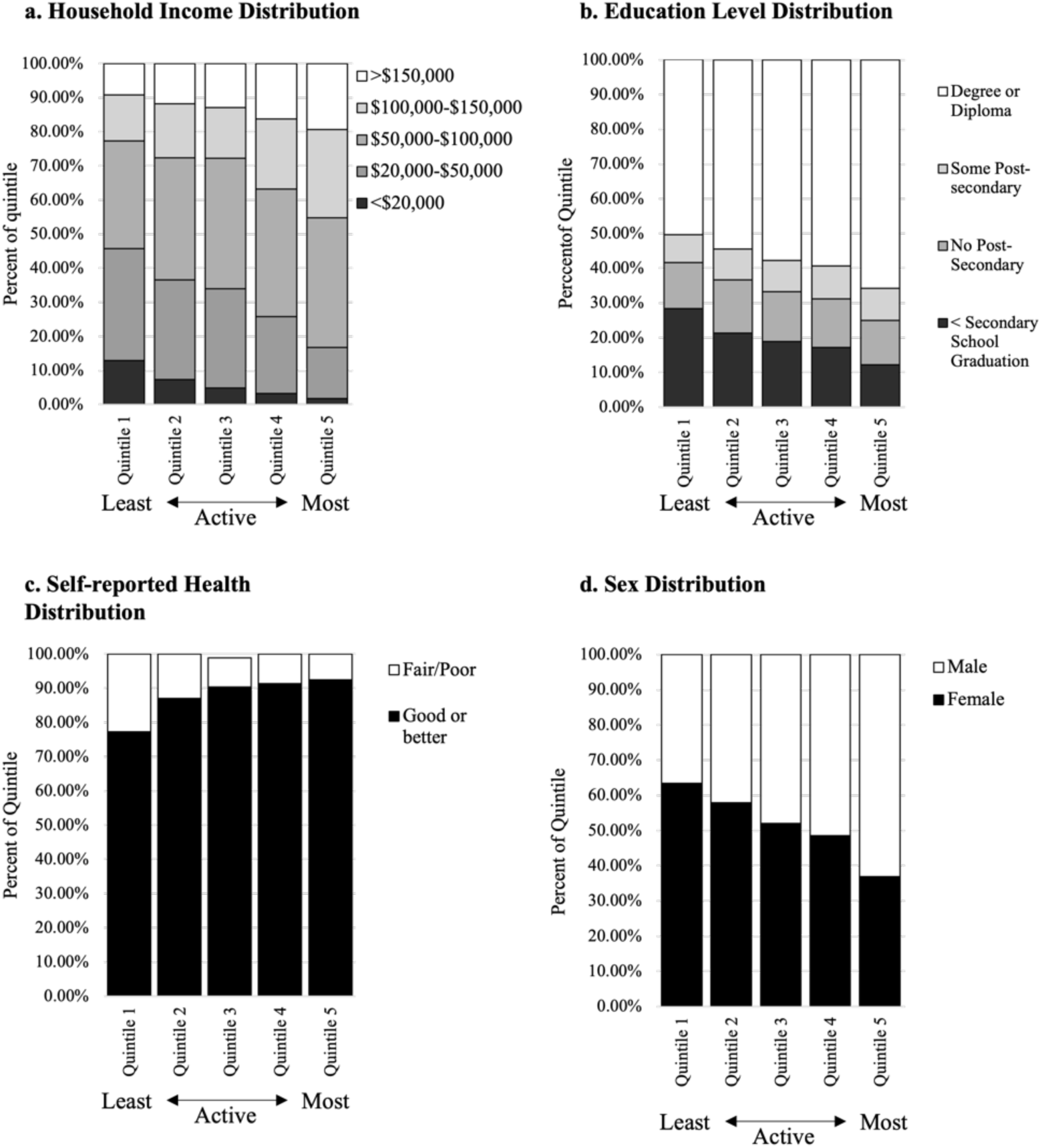
Population Patterns Across PASE Total Score Quintiles. Bar sections represent the percent of people from each outcome present in each PA quintile.

**Table 1.** Weighted Sample Description by Total PASE Score Quintiles.

|  | **1<sup>st</sup> Quintile**<br>20<sup>th</sup><br>Percentile<br>and under<br>(**≤85**) | **2<sup>nd</sup> Quintile**<br>20<sup>th</sup>-40<sup>th</sup><br>Percentiles<br>(**86-121**) | **3<sup>rd</sup> Quintile**<br>40<sup>th</sup>-60<sup>th</sup><br>Percentiles<br>(**122-158**) | **4<sup>th</sup> Quintile**<br>60<sup>th</sup>-80<sup>th</sup><br>Percentiles<br>(**159-217**) | **5<sup>th</sup> Quintile**<br>80<sup>th</sup><br>Percentile<br>and higher<br>(**≥218**) |
| --- | --- | --- | --- | --- | --- |
| PASE Score<br>*Mean (SD)* | 55.42<br>(20.88) | 104.76<br>(10.65) | 139.92<br>(10.56) | 184.21<br>(16.81) | 274.46<br>(49.40) |
| Age<br>*Mean (SD)* | 64.50<br>(11.14) | 62.03<br>(10.22) | 60.65<br>(9.65) | 57.84<br>(8.90) | 53.75<br>(6.90) |
| Sex (% Female) | 1593498<br>(63.41%) | 1405949<br>(57.90%) | 1242232<br>(52.04%) | 1184589<br>(48.53%) | 894034<br>(36.91%) |
| **Household Income** |  |  |  |  |  |
| <\$20,000 | 300,676<br>(12.97%) | 167,649<br>(7.36%) | 109,404<br>(4.82%) | 74,606<br>(3.21%) | 42,569<br>(1.81%) |
| \$20,000-\$50,000 | 760,862<br>(32.83%) | 663,931<br>(29.16%) | 662,613<br>(29.17%) | 526,079<br>(22.65%) | 352,617<br>(15.0%) |
| \$50,000-\$100,000 | 730,957<br>(31.54%) | 815,114<br>(35.80%) | 870,089<br>(38.31%) | 866,227<br>(37.30%) | 893,321<br>(38.01%) |
| \$100,000-\$150,000 | 312,812<br>(13.50%) | 361,940<br>(15.90%) | 337,404<br>(14.86%) | 477,539<br>(20.56%) | 606,031<br>(25.79%) |
| >\$150,000 | 212,228<br>(9.16%) | 267,962<br>(11.77%) | 291,710<br>(12.84%) | 377,923<br>(16.27%) | 455,780<br>(19.39%) |
| **Education** |  |  |  |  |  |
| < Secondary School<br>graduation | 708,806<br>(28.33%) | 518,235<br>(21.38%) | 448,187<br>(18.86%) | 418,294<br>(17.20%) | 294,125<br>(12.17%) |
| No Post-Secondary | 334,207<br>(13.36%) | 371,478<br>(15.33%) | 340,948<br>(14.35%) | 338,868<br>(13.93%) | 310,614<br>(12.85%) |
| Some Post-Secondary | 195,859<br>(7.93%) | 213,665<br>(8.82%) | 215,611<br>(9.07%) | 231,096<br>(9.50%) | 223,671<br>(9.25%) |
| Degree or Diploma | 1,262,905<br>(50.48%) | 1,320,117<br>(54.47%) | 1,371,884<br>(57.72%) | 1,444,050<br>(59.37%) | 1,588,459<br>(65.72%) |
| **Self-reported General Health** |  |  |  |  |  |
| Good or Better | 1,942,178<br>(77.34%) | 2,111,379<br>(87.08%) | 2,154,483<br>(90.34%) | 2,230,005<br>(91.39%) | 2,240,721<br>(92.52%) |
| Fair/Poor | 568,903<br>(22.66%) | 313,177<br>(12.92%) | 230,315<br>(8.61%) | 210,039<br>(8.61%) | 181,265<br>(7.48%) |
| **Diagnosed with Medical Condition (% yes)** |  |  |  |  |  |
| Cancer | 434,035<br>(16.18%) | 331,712<br>(13.67%) | 322,903<br>(13.54%) | 255,873<br>(10.49%) | 206,253<br>(8.53%) |
| Cardiovascular | 1,464,996<br>(16.18%) | 1,120,800<br>(13.67%) | 978,621<br>(13.54%) | 908,399<br>(10.49%) | 697,012<br>(8.53%) |
| Diabetes | 649,577<br>(24.22%) | 401,582<br>(16.56%) | 340,773<br>(14.31%) | 351,987<br>(14.43%) | 287,110<br>(11.88%) |
| Musculoskeletal | 1,641,195<br>(61.57%) | 1,335,815<br>(55.52%) | 1,208,304<br>(51.03%) | 1,180,390<br>(48.64%) | 1,028,833<br>(42.79%) |
| Neurological | 351716<br>(13.21%) | 194,291<br>(8.05%) | 153742<br>(6.47%) | 117121<br>(4.81%) | 73,046<br>(3.02%) |
| Respiratory | 559,938<br>(20.90%) | 442,760<br>(18.30%) | 353508<br>(14.87%) | 309688<br>(12.76%) | 315,664<br>(13.06%) |
| Vision | 1,023,655<br>(38.47%) | 659,128<br>(27.37%) | 544,648<br>(22.98%) | 432,324<br>(17.90%) | 217,394<br>(9.04%) |
| Limitation in one or more ADL/IADL |  |  |  |  |  |
| Yes | 483,496<br>(19.31%) | 160,050<br>(6.59%) | 90,444<br>(3.79%) | 74,213<br>(3.04%) | 71,472<br>(2.95%) |
| No | 2,020,916<br>(80.69%) | 2,267,312<br>(93.41%) | 2,294,273<br>(96.21%) | 2,365,897<br>(96.96%) | 2,348,593<br>(97.05%) |
| Mobility limitation (uses a gait aid) |  |  |  |  |  |
| Yes | 560,878<br>(22.32%) | 259,010<br>(10.67%) | 195,080<br>(8.17%) | 148,558<br>(6.09%) | 111,805<br>(4.62%) |
| No | 1,952,266<br>(77.68%) | 2,169,259<br>(89.33%) | 2,192,079<br>(91.83%) | 2,292,572<br>(93.91%) | 2,310,333<br>(95.38%) |
| Social Support<br>*Mean (SD)* (0-100) | 79.42<br>(19.98) | 82.34<br>(17.43) | 83.58<br>(17.01) | 84.87<br>(15.93) | 84.75<br>(15.84) |
| Social participation – not counting PA |  |  |  |  |  |
| Yearly | 152,251<br>(5.68%) | 105,160<br>(4.34%) | 78,529<br>(3.30%) | 76,207<br>(3.13%) | 79,516<br>(3.28%) |
| Monthly | 650,314<br>(24.25%) | 604,861<br>(24.98%) | 557,435<br>(23.39%) | 519,818<br>(21.33%) | 612,896<br>(25.31%) |
| Weekly | 1,627,427<br>(60.67%) | 1,500,372<br>(61.97%) | 1,500,824<br>(62.98%) | 1,583,489<br>(64.98%) | 1,467,522<br>(60.60%) |
| Daily | 211,669<br>(7.89%) | 197,278<br>(8.15%) | 242,879<br>(10.19%) | 252,416<br>(10.36%) | 261,194<br>(10.79%) |
| Self-perceived Social Status |  |  |  |  |  |
| 1-2 (low) | 370,695<br>(14.77%) | 220,142<br>(9.52%) | 203,946<br>(8.81%) | 171,915<br>(7.25%) | 140,907<br>(5.97%) |
| 3-4 | 360,737<br>(14.38%) | 305,961<br>(13.24%) | 274,750<br>(11.87%) | 262,971<br>(11.09%) | 256,707<br>(10.88%) |
| 5-6 | 931,678<br>(37.13%) | 913,536<br>(39.52%) | 906,922<br>(39.18%) | 940,728<br>(39.68%) | 844,197<br>(35.77%) |
| 7-8 | 709,631<br>(28.28%) | 763,930<br>(33.05%) | 811,798<br>(35.07%) | 863,360<br>(36.42%) | 980,970<br>(41.57%) |
| 9-10 (high) | 136,319<br>(5.43%) | 108,174<br>(4.68%) | 117,156<br>(5.06%) | 131,887<br>(5.56%) | 136,985<br>(5.81%) |
| Rural | 420,900<br>(16.75%) | 549,574<br>(22.63%) | 538,361<br>(22.55%) | 523,659<br>(21.45%) | 570,950<br>(23.57%) |
| Urban | 2,092,244<br>(83.25%) | 1,878,696<br>(77.37%) | 1,848,798<br>(77.45%) | 1,917,470<br>(78.55%) | 1,851,189<br>(76.43%) |

## DISCUSSION

This is the first paper to describe usual PA by type of activity and amount in Canadians 45-85 years old across all 10 provinces. This descriptive analysis demonstrated the relationships between several social determinants of health and PA behaviours. Notably, these relationships were most pronounced in those from the lowest socioeconomic groups compared to all other subgroups. Targeted PA promotion efforts appear warranted based on the heterogeneity of PA behaviours by age, income level, and region in middle-aged and older Canadians. While several factors appeared to have salient relationships with PA levels, future research is needed to determine the directionality of these relationships.

Describing Canadians by their total PA levels demonstrated a clear gap in several social determinants of health between the most active (top 20% of PASE scores Q5) and the least active (bottom 20% of PASE scores Q1). Individuals in the bottom 20% of PA were more likely to be female, have lower household income and education levels, and report worse health. A similar intersectionality was seen in two population-based analyses of adults (18+ years) meeting recommended levels of MVPA in Brazil (n=58,429)^22^ and Canada (n=15,510).^13^ Mielke et al. (2022) found that 48.0% of white males in Brazil from the highest income quartile with a university degree, compared to only 9.0% of non-white females with low education and income met recommended MVPA levels.^22^ Colley et al. (2023), showed similar patterns using latent class analysis of the Canadian Health Measures Survey, where individuals least likely to meet guidelines were more likely to be female, older age, and have lower income and education.^13^ These disparities in PA participation are concerning and suggest further investigation into the determinants of PA behaviour in these individuals is needed, and may improve interventions for these populations.

The exploration of socioeconomic factors showed a noteworthy distinction for the lowest levels. For both education and household income, the lowest categories demonstrated the most considerable differences in mean PASE scores compared to other categories. This could indicate the presence of a threshold for these two factors where a greater negative effect on PA is seen up to a certain point (i.e., secondary school graduation and $20,000+). The relationship with household income appeared to be attenuated by age but did not account for the overall trend of increasing PA with household income. Interventions and policies targeting PA promotion among individuals from lower income and education groups is of particular importance for improving population health in Canada.

Being female and older were related to lower PA levels as shown by previous work; however, our findings highlight the types of activities favoured by these groups that could be targeted as part of PA promotion efforts. For example, the lower participation in heavier housework reported among females could be a target for PA promotion. Another trend that may be unique to Canada and countries with similar climates was the presence of high PA throughout the colder months in certain regions of Canada. The PA patterns in British Columbia and the Prairies contrast those reported in previous literature and from Central and Atlantic Canada.^14^ Regardless of the season, Atlantic Canada appears less active overall and may benefit from specific PA promotion efforts in addition to national messaging.

Finally, while several variables exhibited relationships with PA behaviours, including the presence of mobility and ADL limitations and certain chronic conditions, this analysis was cross-sectional. Therefore, we cannot infer the directionality of these relationships and future work exploring longitudinal associations of these variables is warranted. There are several other limitations to consider when interpreting the data from this study. Firstly, the PASE was designed for older adults (65+ years) and has not been validated among middle-aged adults. However, the questionnaire was designed to be a comprehensive measure of PA in later life as it captures both light-intensity PA and work-related activities. Finally, the CLSA sample only represents community-dwelling Canadians from the 10 provinces (i.e., not the territories) and excludes persons living in Federal and other provincial First Nations settlements and full-time Canadian armed forces. However, the extensive dataset of the CLSA allowed for a detailed description of population characteristics, which should be considered before generalizing these results.

## CONCLUSION

Middle-aged and older Canadians’ PA behaviour is heterogeneous in terms of both the amount and types of PA completed. Social determinants of health clearly influence PA levels in Canada, and targeted interventions and promotion efforts are warranted. Noticeable relationships between PA behaviours and health outcomes, including limitations in mobility and ADLs and chronic conditions, warrant further investigation via longitudinal analyses to determine directionality.

## AUTHOR CONTRIBUTIONS

CD and MKB developed the research question and initial methods. CD conducted analysis with feedback and support from MKB, LEG, and JR. All authors contributed to the interpretation of results and manuscript revisions.

## ACKNOWLEDGEMENTS

This research was made possible using the data/biospecimens collected by the Canadian Longitudinal Study on Aging (CLSA). Funding for the Canadian Longitudinal Study on Aging (CLSA) is provided by the Government of Canada through the Canadian Institutes of Health Research (CIHR) under grant reference: LSA 94473 and the Canada Foundation for Innovation, as well as the following provinces, Newfoundland, Nova Scotia, Quebec, Ontario, Manitoba, Alberta, and British Columbia. This research has been conducted using the CLSA dataset Baseline Tracking Dataset Version 4.0, Baseline Comprehensive Dataset Version 7.0, under Application Number 190233. The CLSA is led by Drs. Parminder Raina, Christina Wolfson and Susan Kirkland. NDVI metrics, indexed to DMTI Spatial Inc. postal codes, were provided by CANUE (Canadian Urban Environmental Health Research Consortium). The opinions expressed in this manuscript are the author’s own and do not reflect the views of the Canadian Longitudinal Study on Aging.

## DATA AVAILABILITY

Data are available from the Canadian Longitudinal Study on Aging (www.clsa-elcv.ca) for researchers who meet the criteria for access to de-identified CLSA data.

## Supplemental Material

## Appendix A. CLSA sample and recruitment

The CLSA excluded individuals living in long-term or institutional care settings at the time of recruitment, full-time members of the Canadian armed forces, persons living on federal First Nations reserves or provincial First Nations settlements, and residents of the Canadian territories and some remote regions. The CLSA’s sample is divided into two cohorts, the tracking cohort (21,241) which uses assessments completed by telephone interviews and the comprehensive cohort (30,097) which consist of in-person (in-home and at data collection site) and telephone assessments. Due to the nature of the comprehensive cohort assessments participants in this group were recruited from a 25-50km range around one of the 11 data collection sites located in seven provinces. Further details on the recruitment and sampling methods of the CLSA can be found elsewhere. As the purpose of this work was to describe and generalize to the target population, other than completing the maintaining contact questionnaire, no further exclusion criteria were applied.

More details on sampling and recruitment are available here:

Raina P, Wolfson C, Kirkland S, Griffith LE, Balion C, Cossette B, et al. Cohort profile: The Canadian Longitudinal Study on Aging (CLSA). Int J Epidemiol. 2019;48(6):1752-3j.
Raina PS, Wolfson C, Kirkland SA, Griffith LE, Oremus M, Patterson C, et al. The Canadian longitudinal study on aging (CLSA). Can J Aging. 2009;28(3):221-9.

## Appendix B. Physical Activity Scale for the Elderly and PA outcomes

### Physical activity outcomes

#### 1. Total Physical Activity

The total PASE score is calculated based on question responses and the developed activity weights.^12^ The score ranges from 0 to over 400, with higher scores representing greater PA levels. This outcome has shown acceptable test-retest reliability and construct validity (e.g., walking, physical function, general health).^14^ The PASE total score was calculated for all participants with the required information available, rounded to a whole number as per the PASE instruction manual.^15^

#### 3. Prevalence of activity types

The percentage of participants who completed each activity in the previous 7-days was calculated.

#### 4. 150-minutes of MVPA guidelines

The total time spent in walking, moderate recreation, strenuous recreation, and exercise were calculated using the methods by Mayo et al. (2021), details below.^16^ Individuals with at least 150-minutes of MVPA in the last 7-days were said to have met the guideline.

To arrive at the total number of minutes per week of MVPA we first calculated time spent in walking, moderate recreation, strenuous recreation, and exercise activities. Following the methods used by Mayo et al. in their 2021 analysis a median value was created for each frequency and duration range (see table B1.). Except for the final duration, which does not provide an upper limit, in this case this most conservative estimate was taken (i.e., 4 hours). The amount of time was converted from hours a day to minutes and then multiped by the frequency. After the minutes per week for each of the four available activities was calculated and summed, a dichotomous outcome was derived.

Reference:

Mayo A, Sénéchal M, Boudreau J, Bélanger M, Bouchard DR. Potential functional benefits of a comprehensive evaluation of physical activity for aging adults: a CLSA cross-sectional analysis. Aging Clin Exp Res. 2021;33(2):285-9.

**Table B1.**
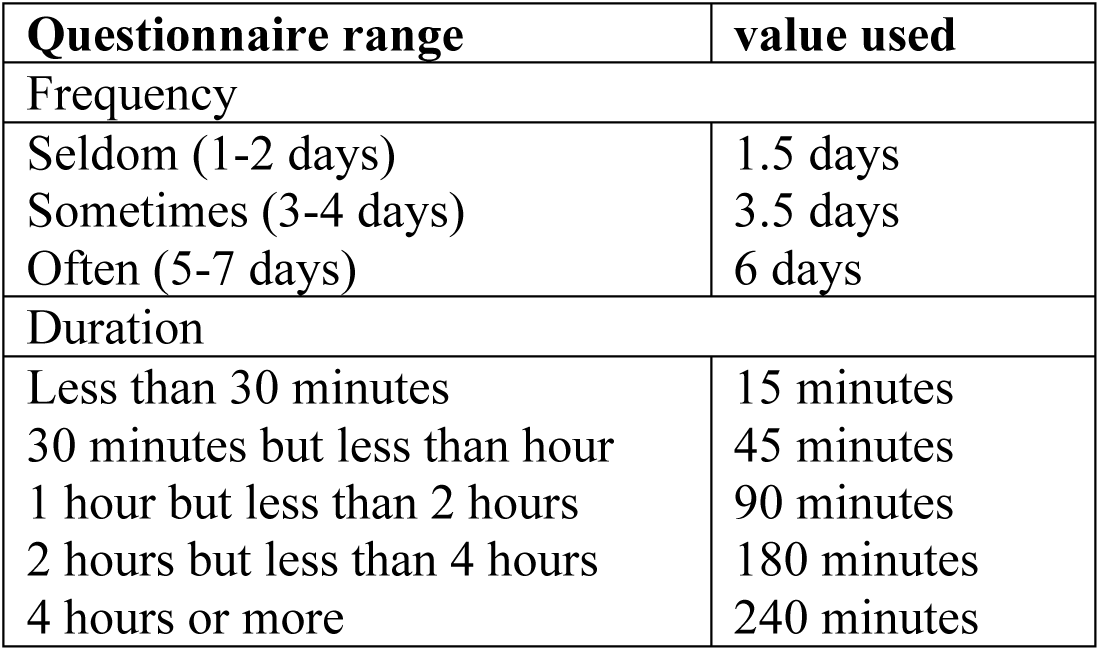
150-minute outcome operationalization.

**Table B2.**
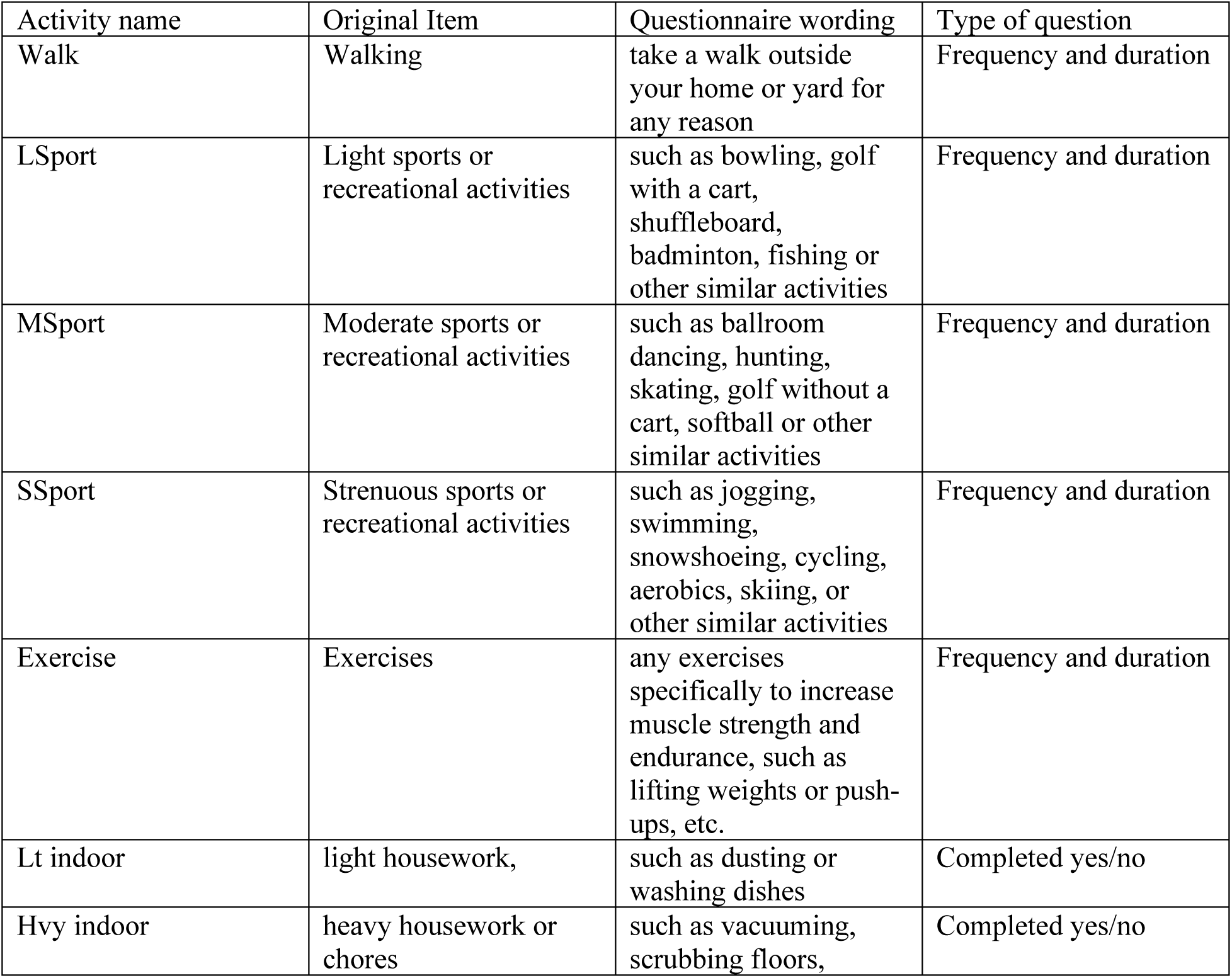
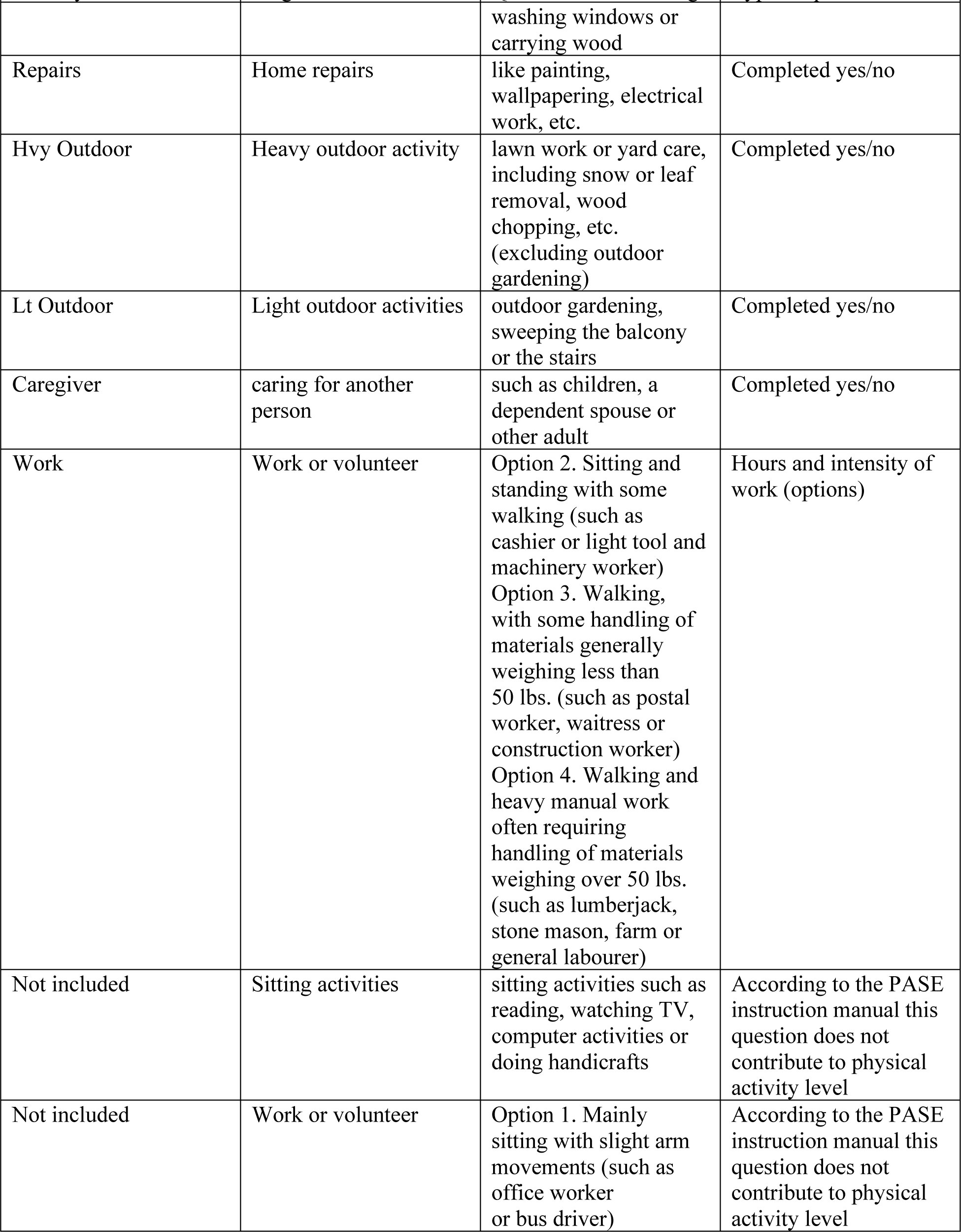
PASE Items and Questions.

| Activity name | Original Item | Questionnaire wording | Type of question |
| --- | --- | --- | --- |
| Walk | Walking | take a walk outside your home or yard for any reason | Frequency and duration |
| LSport | Light sports or recreational activities | such as bowling, golf with a cart, shuffleboard, badminton, fishing or other similar activities | Frequency and duration |
| MSport | Moderate sports or recreational activities | such as ballroom dancing, hunting, skating, golf without a cart, softball or other similar activities | Frequency and duration |
| SSport | Strenuous sports or recreational activities | such as jogging, swimming, snowshoeing, cycling, aerobics, skiing, or other similar activities | Frequency and duration |
| Exercise | Exercises | any exercises specifically to increase muscle strength and endurance, such as lifting weights or push-ups, etc. | Frequency and duration |
| Lt indoor | light housework, | such as dusting or washing dishes | Completed yes/no |
| Hvy indoor | heavy housework or chores | such as vacuuming, scrubbing floors, washing windows or carrying wood | Completed yes/no |
| Repairs | Home repairs | like painting, wallpapering, electrical work, etc. | Completed yes/no |
| Hvy Outdoor | Heavy outdoor activity | lawn work or yard care, including snow or leaf removal, wood chopping, etc. (excluding outdoor gardening) | Completed yes/no |
| Lt Outdoor | Light outdoor activities | outdoor gardening, sweeping the balcony or the stairs | Completed yes/no |
| Caregiver | caring for another person | such as children, a dependent spouse or other adult | Completed yes/no |
| Work | Work or volunteer | Option 2. Sitting and standing with some walking (such as cashier or light tool and machinery worker)<br>Option 3. Walking, with some handling of materials generally weighing less than 50 lbs. (such as postal worker, waitress or construction worker)<br>Option 4. Walking and heavy manual work often requiring handling of materials weighing over 50 lbs. (such as lumberjack, stone mason, farm or general labourer) | Hours and intensity of work (options) |
| Not included | Sitting activities | sitting activities such as reading, watching TV, computer activities or doing handicrafts | According to the PASE instruction manual this question does not contribute to physical activity level |
| Not included | Work or volunteer | Option 1. Mainly sitting with slight arm movements (such as office worker or bus driver) | According to the PASE instruction manual this question does not contribute to physical activity level |

## Appendix C. Subgroup stratifications

**Table C1.** Subgroup stratification details.

| Correlate(s) for subgroup | Number of groups | Details |
| --- | --- | --- |
| Age and sex | 8 | Four age groups (45-54, 55-64, 65-74, 75+) for both males and females |
| Household income | 5 | <\$20,000,<br>\$20,000-<\$50,000,<br>\$50,000-<\$100,000, \$100,000-<\$150,000, and<br>\$150,000+ |
| Household income and age | 20 | Four age groups and five income levels |
| Education | 4 | Less than secondary school graduation,<br>No post-secondary education (but graduated from<br>secondary school),<br>Some post-secondary education, and<br>Received a degree/diploma |
| Material Deprivation | 5 | Based on the Material and Social Deprivation index which describes the geographical variation in deprivation (six socio-economic indicators) was used to explore the two domains of deprivation, material and social.<sup>1,21, 2</sup> Subgroups were based on quintiles representing those with the most privilege (Q1) to the most deprived (Q5) |
| Social Deprivation | 5 |  |
| Region and season | 16 | four geographical regions (British Columbia, Prairies, Central, and Atlantic) and four seasons (Jan-Mar, Apr-Jun, Jul-Sep, Oct-Dec)<sup>3</sup> |

## Appendix D. Sample, target population and missing variable description

**Table D1.** Sample and Target Population Descriptors.

| Variable | Weighted<br>(target population) | Unweighted<br>(sample) |
| --- | --- | --- |
| **Complete MCQ** | 12,365,513 | 47,840 |
| **Age mean(SD)** | 59.783 (SD 10.181) | 62.98 (SD 10.35) |
| **Sex (Female)** | 6,406,677 (51.81%) | 24,391 (50.98%) |
| **Cultural/racial background** |  |  |
| European | 11,735,186 (95.01%) | 45,533 (96.20%) |
| Non-European | 371,511 (3.94%) | 1,232 (2.60%) |
| Multiple origins | 129,738 (1.05%) | 567 (1.20%) |
| **Had a fall in the previous 12 months** |  |  |
| Yes | 1,373,926 (11.12%) | 5,449 (11.40%) |
| **Diagnosed with a medical condition (Yes)** |  |  |
| Cancer | 1,550,777 (12.56%) | 7,301 (18.05%) |
| Cardiovascular | 5,274,612 (42.90%) | 21,809 (45.91%) |
| Diabetes | 2,031,030 (16.45%) | 8,125 (17.02%) |
| Musculoskeletal | 6,394,538 (52.11%) | 25,440 (53.77%) |
| Neurological | 889,916 (7.23%) | 3,618 (7.62%) |
| Respiratory | 1,981,558 (16.08%) | 7,735 (16.24%) |
| **Number of medications** |  |  |
| None | 2,924,394 (23.78%) | 8,429 (17.67%) |
| One | 2,140,529 (17.41%) | 6,863 (14.39%) |
| Two | 1,721,843 (14.00%) | 6,050 (12.68%) |
| 3+ | 5,510,482 (44.81%) | 26,361 (55.26%) |
| **Self-reported general health** |  |  |
| Good or better | 10,833,305 (87.68%) | 42,879 (89.70%) |
| Fair/poor | 1,522,737 (12.32%) | 4,922 (10.30%) |
| **Education** |  |  |
| < secondary school graduation | 2435789 (19.77%) | 3,152 (6.61%) |
| No post-secondary education | 1718823 (13.95%) | 5,182 (10.86%) |
| Some post-secondary education | 1093640 (8.88%) | 3,570 (7.48%) |
| Post-secondary degree/diploma | 7074034 (57.41%) | 35,815 (75.05%) |
| **Household income** |  |  |
| less than \$20,000 | 709,763 (6.07%) | 2,513 (5.61%) |
| \$20,000 or more, but less than \$50,000 | 3,027,389 (25.88%) | 11,096 (24.77%) |
| \$50,000 or more, but less than \$100,000 | 4,225,053 (36.11%) | 16,116 (35.98%) |
| \$100,000 or more, but less than \$150,000 | 2,117,014 (18.19%) | 8,300 (18.53%) |
| \$150,000 or more | 1,619,917 (13.85%) | 6,765 (15.01%) |
| **Type of dwelling** |  |  |
| House (single detached, semi-detached, duplex or townhouse) | 10,540,348 (85.26%) | 39,153 (81.87%) |
| Apartment or condominium | 1,629,504 (13.18%) | 8,029 (16.79%) |
| Other | 193,315 (1.56%) | 642 (1.34%) |
| **Marital status** |  |  |
| In a relationship | 9,296,749 (75.18%) | 32,444 (68.86%) |
| Widowed | 947,741 (7.66%) | 4,999 (10.61%) |
| Divorced/separated | 1,152,216 (9.32%) | 6,001 (12.74%) |
| Single | 793,693 (6.42%) | 3,675 (7.80%) |
| **Social support** (MOS Scale) | 82.98 (17.44) | 81.93 (17.39) |
| Functional Social Support (0-100) |  |  |
| **Total PASE score** mean (SD) | 151.094 (SD 79.11) | 141.53 (SD75.17) |
| **Completed 150 minutes of MVPA** | 7,959,951 (64.80%) | 32,152 (67.60%) |
| **PA represent last 12 months** |  |  |
| Agreed | 8,691,980 (70.29%) | 3,365,512 |
| Neither agree nor disagree | 250,285 (2.02%) | (68.82%) |
| Disagreed | 3,365,512 (27.22%) | 1,004 (2.11%) |
|  |  | 13,836 (29.07%) |

**Table D2.** Number of missing values for variables in aim 1 analyses.

|  | Stratification |  |  |  |  |  |  |  |  |
| --- | --- | --- | --- | --- | --- | --- | --- | --- | --- |
|  | Age | Sex | Household income | Education | Material deprivation scale | Social deprivation scale | Region | Season | Retirement status |
| Total PASE Score/Proportion of score | 633 | 633 | 3621 | 751 | 2642 | 2642 | 633 | 633 | 447 |
| 150 minutes of MVPA | 275 | 275 | 3295 | 549 | 2278 | 2278 | 275 | 275 | 447 |
| Walk | 69 | 69 | 3112 | 190 | 2076 | 2076 | 69 | 69 | 241 |
| Light recreation | 22 | 22 | 3070 | 143 | 2030 | 2030 | 22 | 22 | 194 |
| Moderate recreation | 25 | 25 | 3070 | 146 | 2033 | 2033 | 25 | 25 | 197 |
| Strenuous recreation | 28 | 28 | 3073 | 149 | 2036 | 2036 | 28 | 28 | 200 |
| Exercise | 42 | 42 | 3086 | 163 | 2048 | 2048 | 42 | 42 | 214 |
| Light indoor housework | 17 | 17 | 3065 | 138 | 2026 | 2026 | 17 | 17 | 189 |
| Heavy indoor housework | 21 | 21 | 3068 | 142 | 2030 | 2030 | 21 | 21 | 193 |
| Light outdoor housework | 15 | 15 | 3061 | 136 | 2024 | 2024 | 15 | 15 | 187 |
| Heavy outdoor housework | 19 | 19 | 3066 | 140 | 2028 | 2028 | 19 | 19 | 191 |
| Caregiving | 36 | 36 | 3082 | 157 | 2043 | 2043 | 36 | 36 | 208 |
| Home repairs | 36 | 36 | 3084 | 157 | 2044 | 2044 | 36 | 36 | 208 |
| Work or volunteer | 166 | 166 | 3205 | 286 | 2166 | 2166 | 166 | 166 | 338 |

**Table D3.** Number of missing values for variables in aim 2 analyses.

|  | Outcome |  |
| --- | --- | --- |
|  | None | Total PA Quintile |
| Age | 0 | 633 |
| Sex | 0 | 633 |
| Household income | 3050 | 3621 |
| Education | 121 | 751 |
| Cancer | 99 | 732 |
| Cardiovascular | 334 | 967 |
| Diabetes | 116 | 749 |
| Musculoskeletal | 530 | 1149 |
| Neurological | 331 | 956 |
| Respiratory | 217 | 846 |
| Vision | 728 | 1351 |
| Self-reported general health | 39 | 671 |
| ADL/IADL Limitation | 55 | 687 |
| Mobility limitation | 0 | 633 |
| Social participation | 0 | 633 |
| Perceived social standing | 0 | 633 |
| Social support | 1562 | 2145 |
| Rural vs Urban | 0 | 633 |

## Appendix E. Age and sex stratification

**Figure E1.**
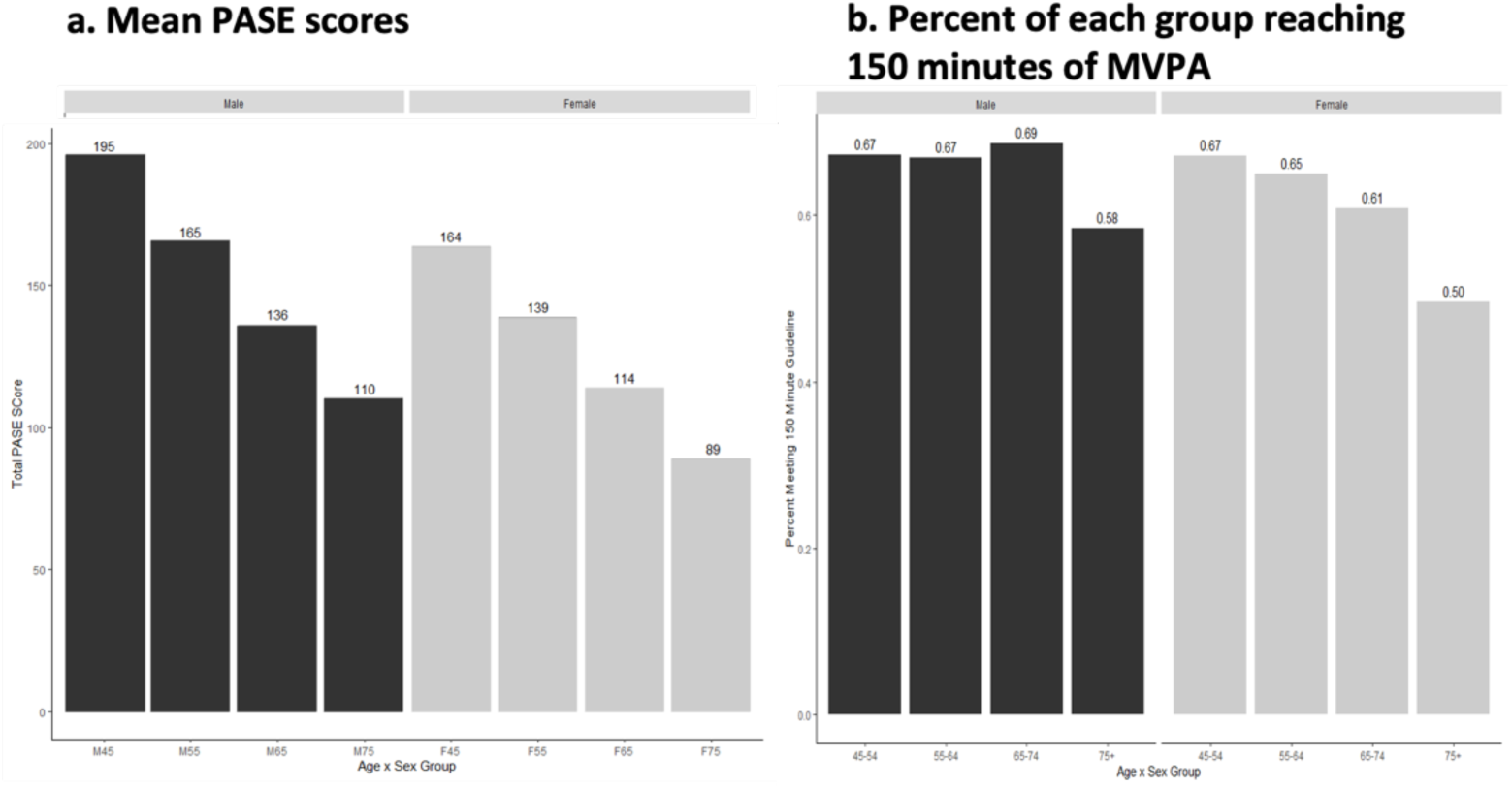
Amount of physical activity sex and age stratification.

**Figure E2.**
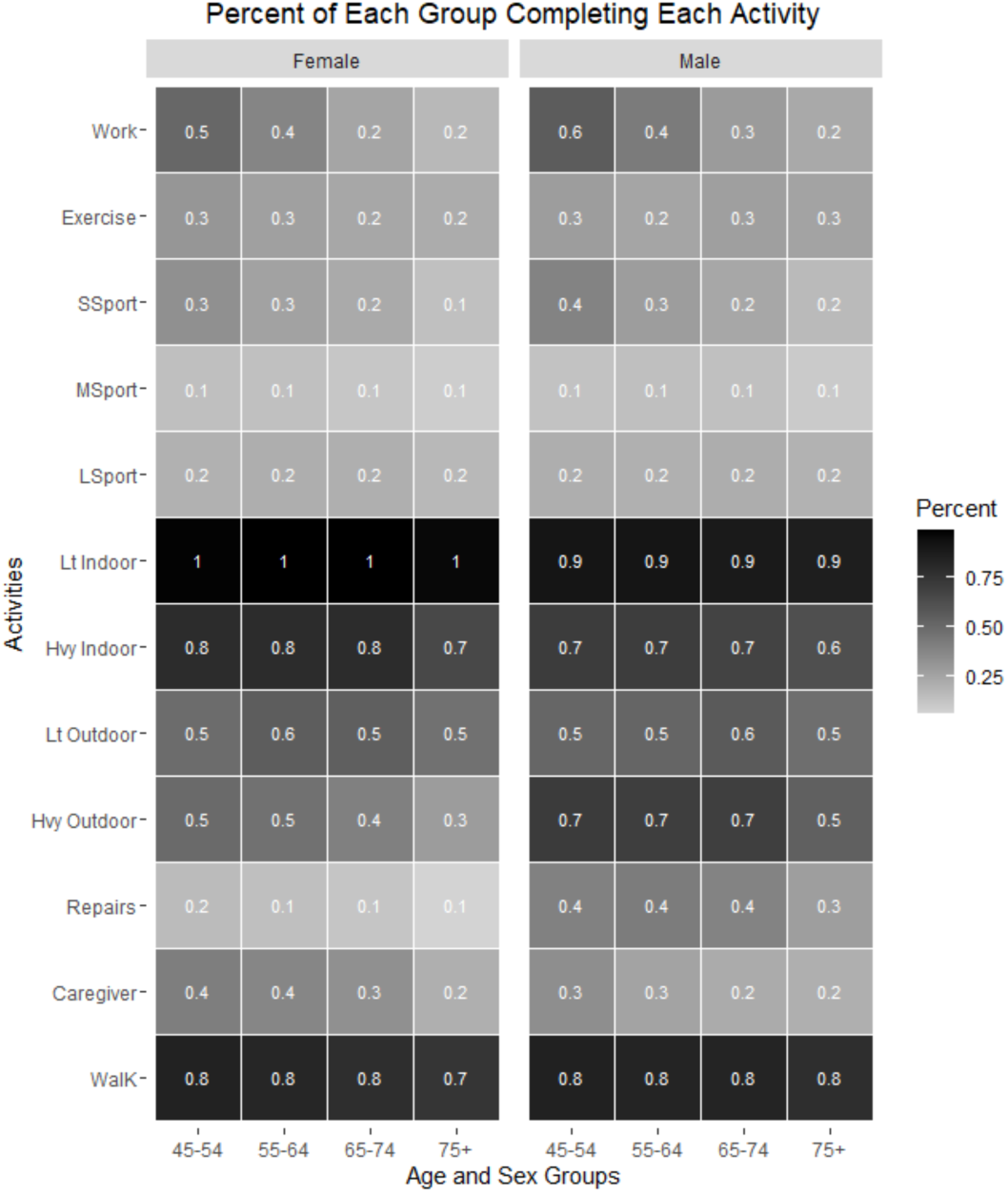
Percentage of each sex and age group participating in activity types.

## Appendix F. Household Income Stratification

**Figure F1.**
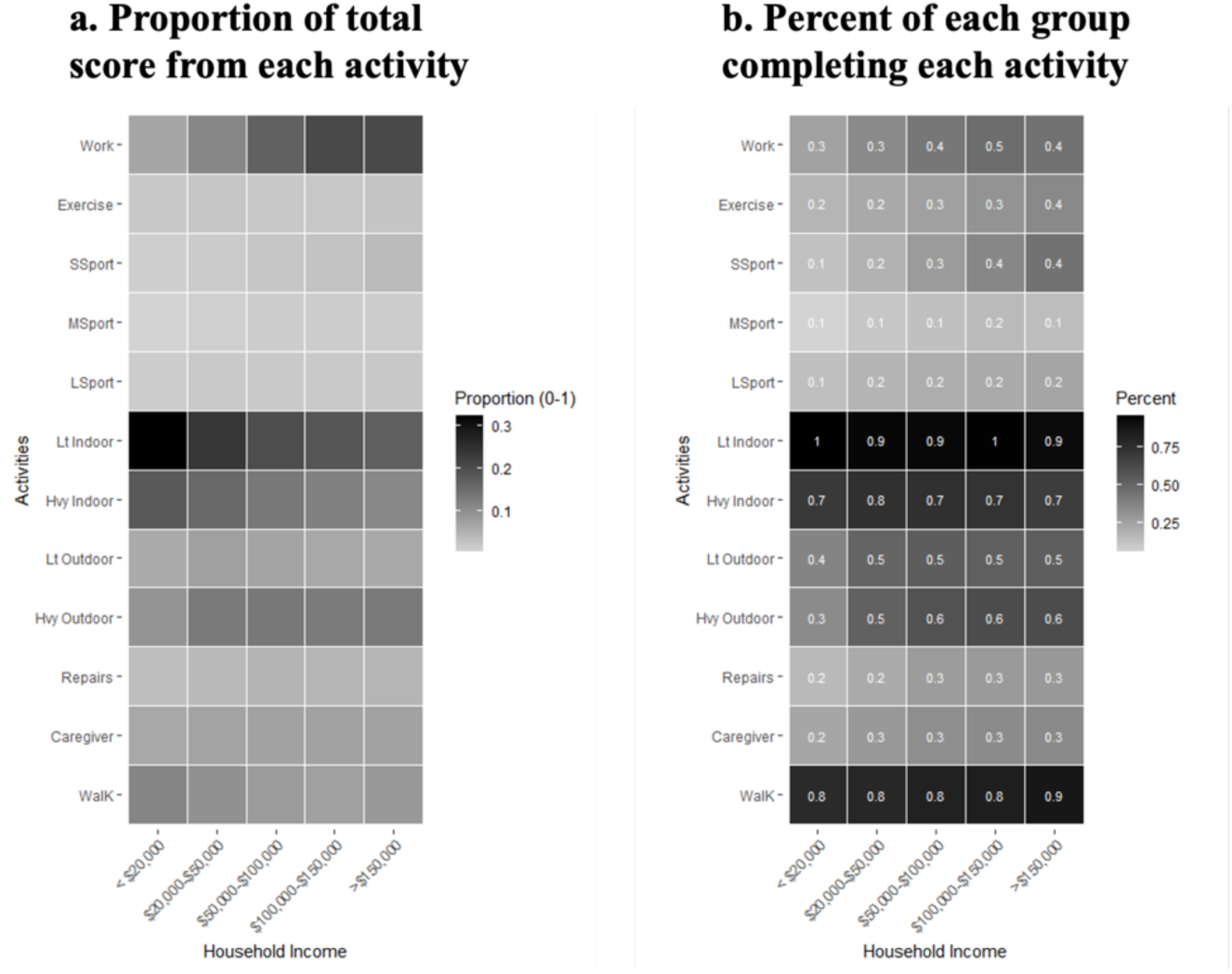
Activity type heat maps for household income.

**Figure F2.**
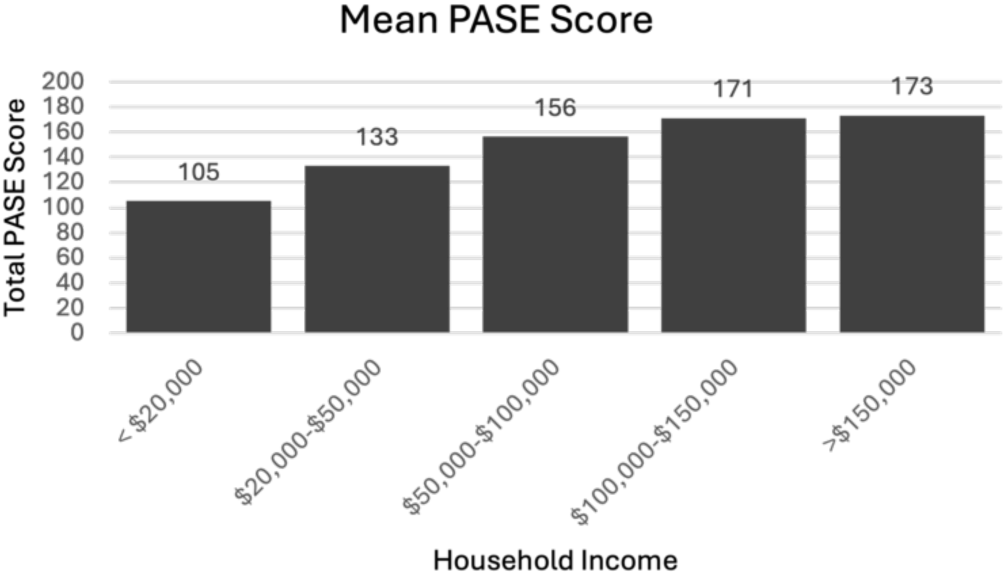
Mean PASE scores by household income.

**Figure F3.**
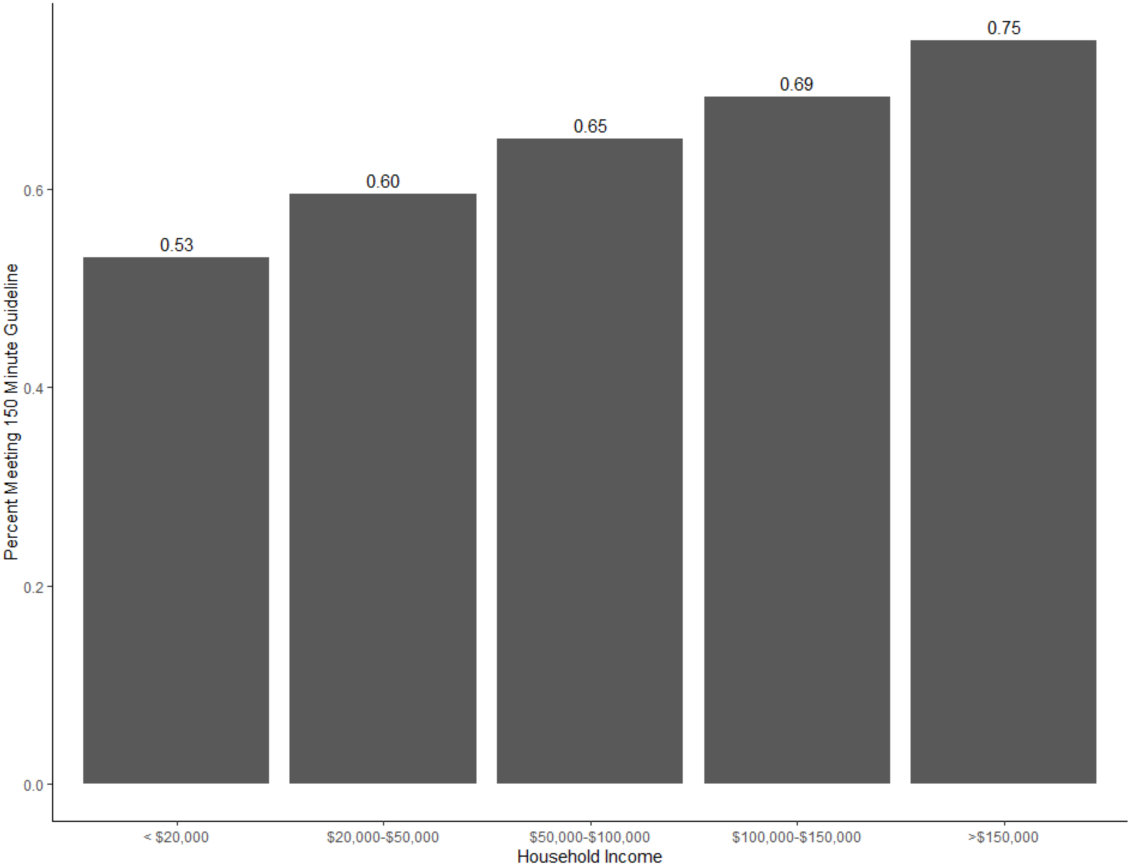
Precent of target population completing 150 minutes of MVPA by household income.

**Figure F4.**
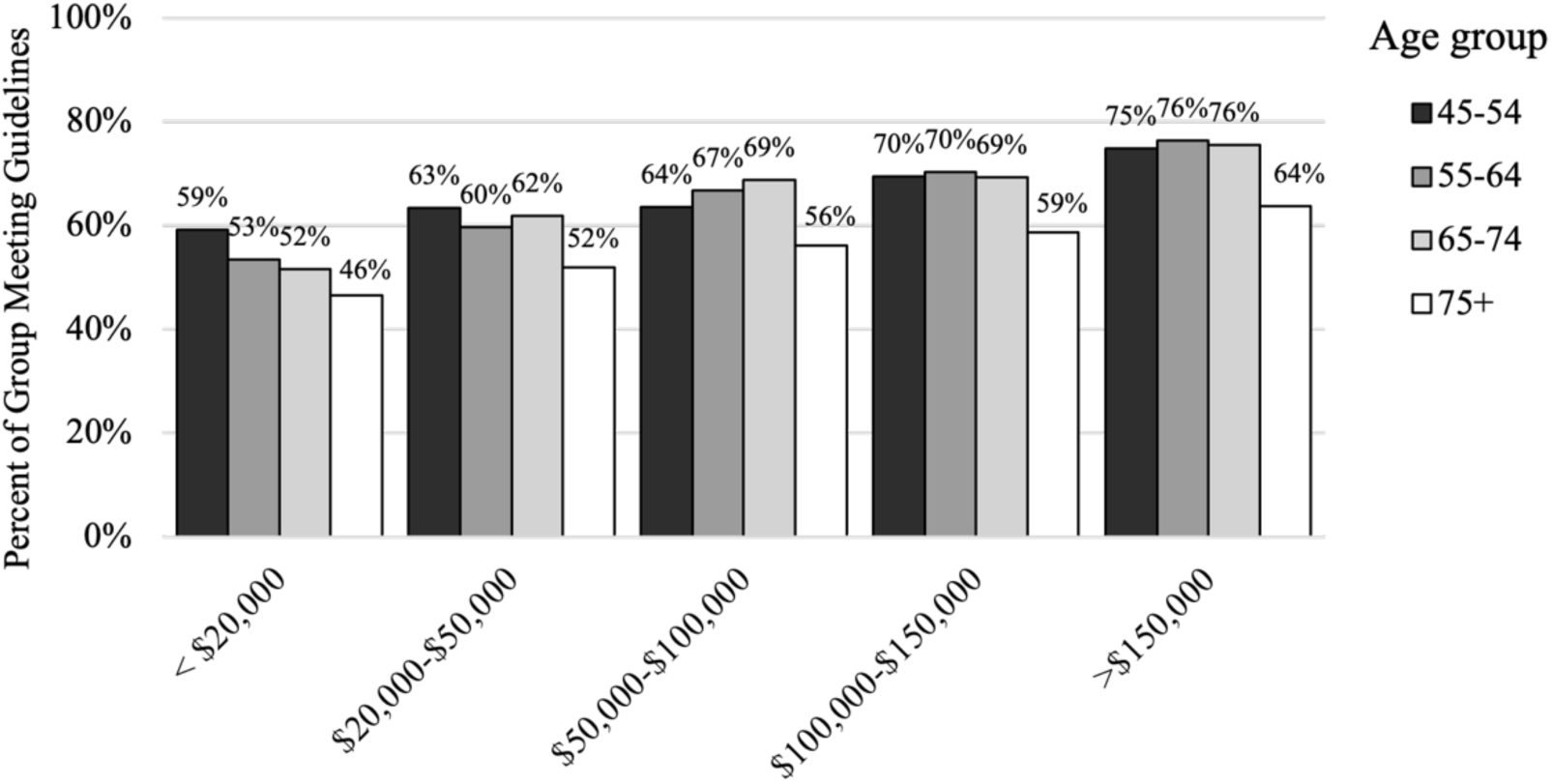
Percent of target population completing 150 minutes of MVPA: Age and income.

**Figure F5.**
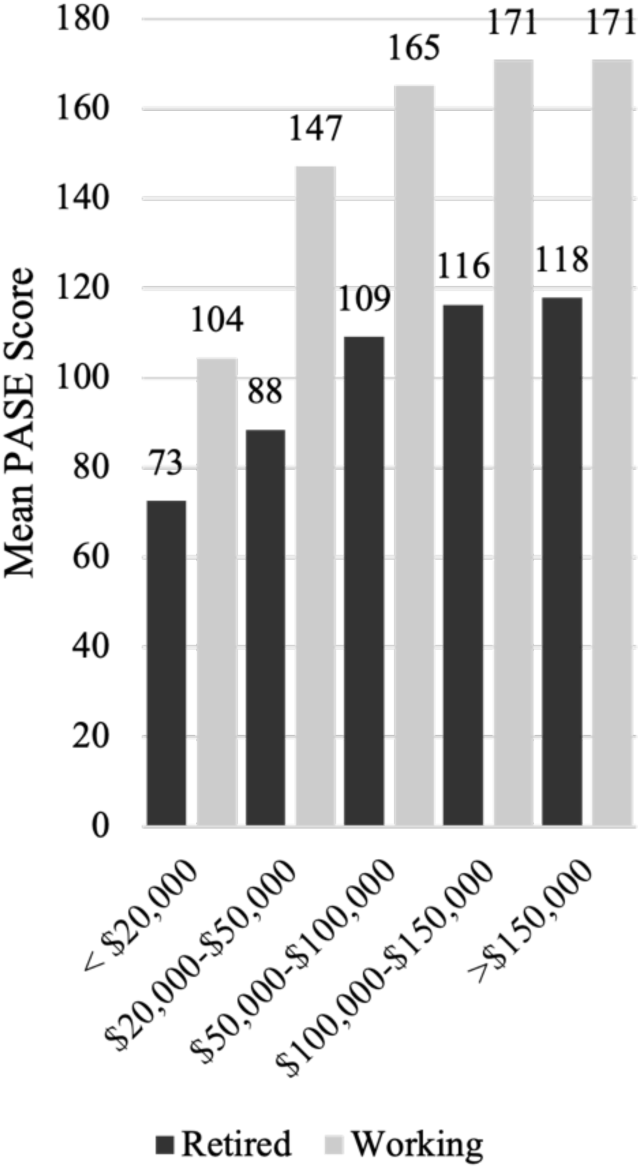
Mean PASE scores comparing retirement status across household income levels.

## Appendix G. Education Level Stratification

**Figure G1.**
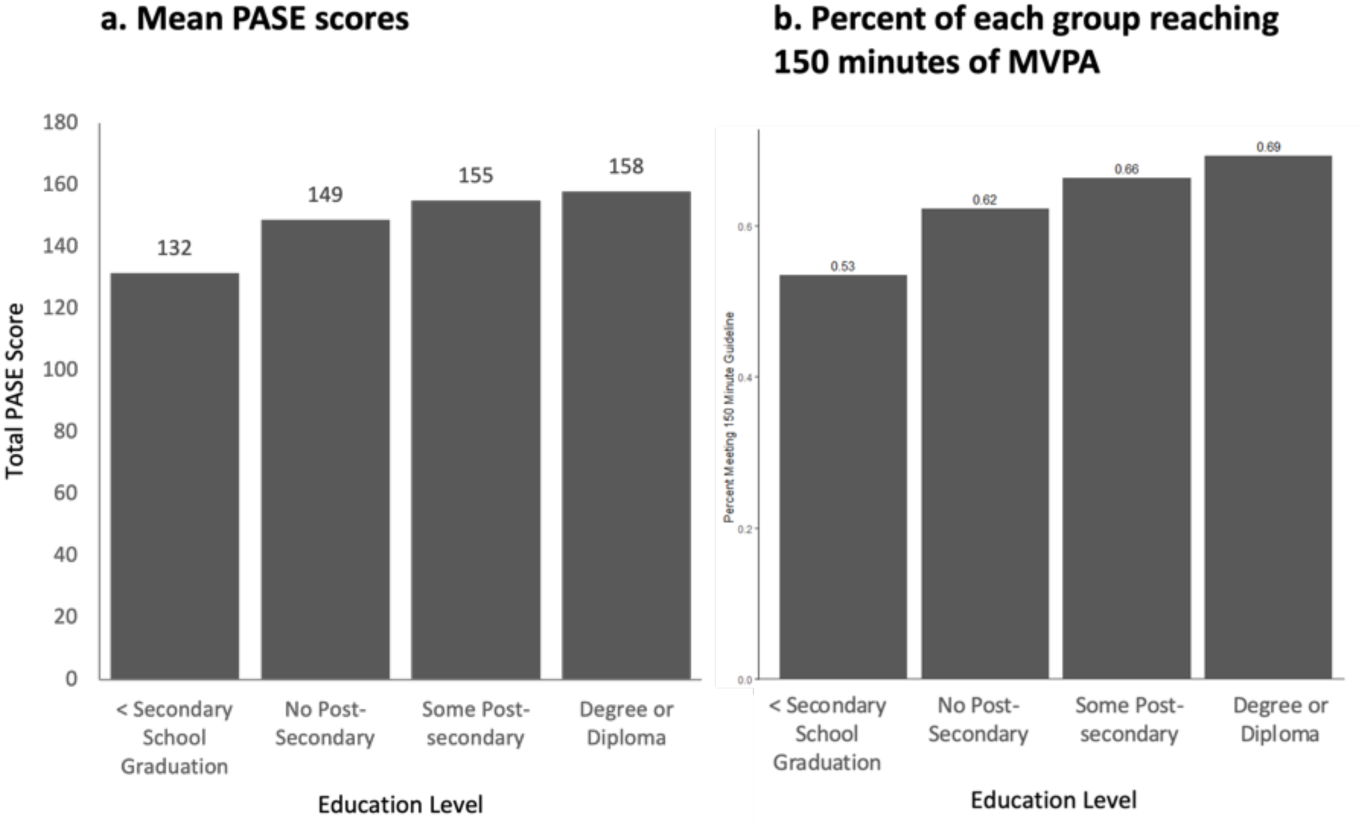
Amount of physical activity by education levels.

**Figure G2.**
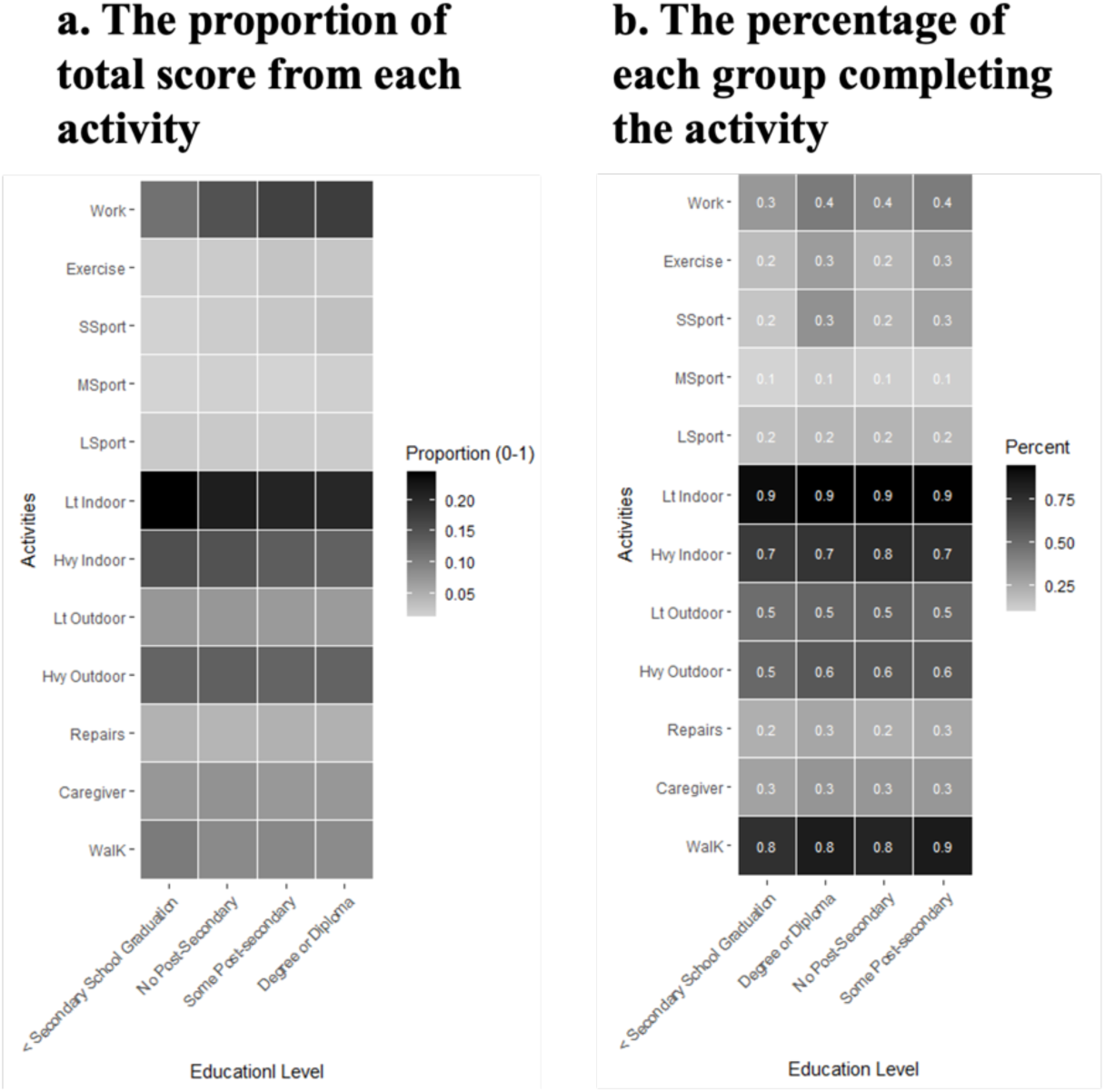
Activity type heat maps by education level.

## Appendix H. Material and Social Deprivation Stratification

**Figure H1.**
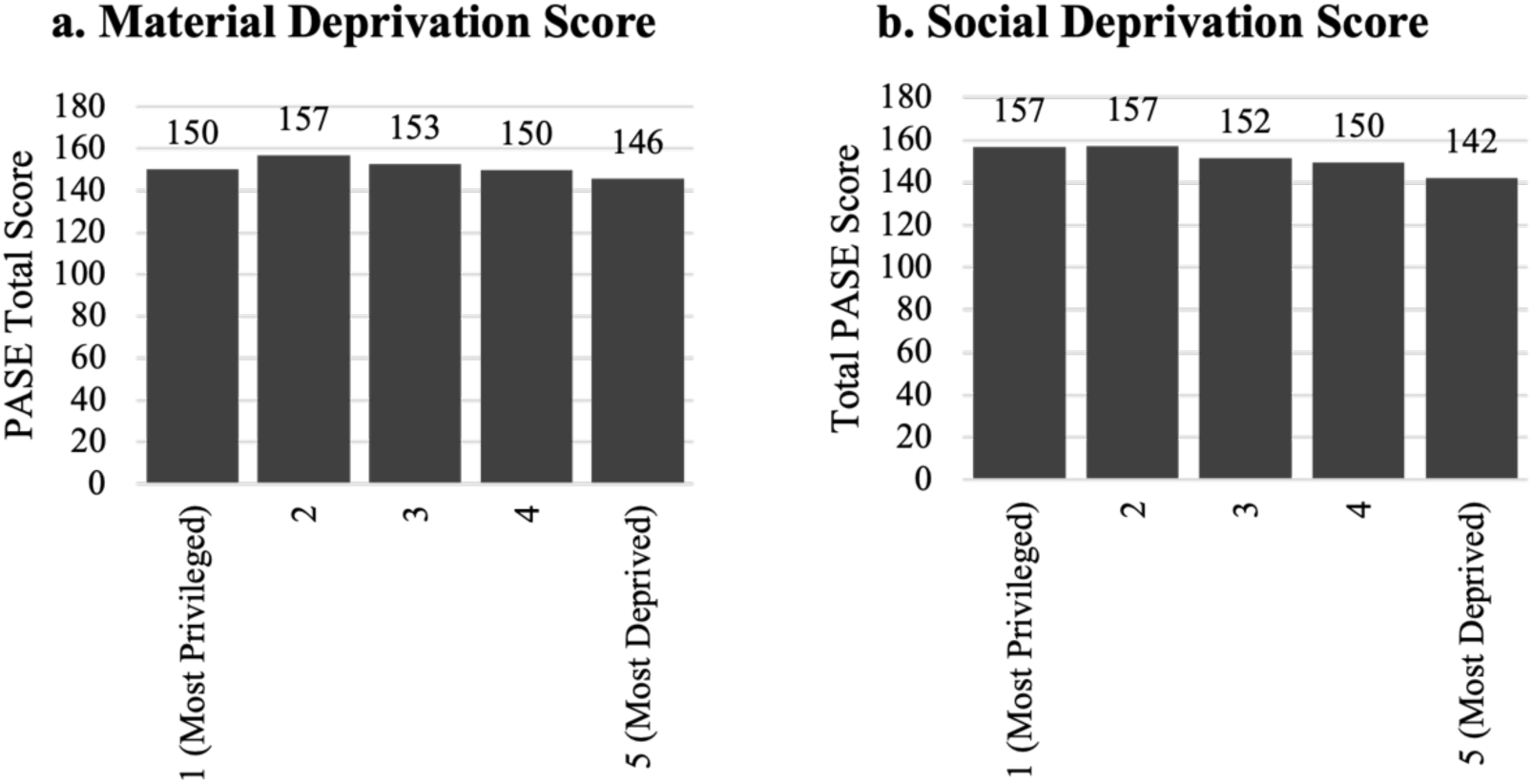
Mean PASE scores for material and social deprivation quintiles.

**Figure H2.**
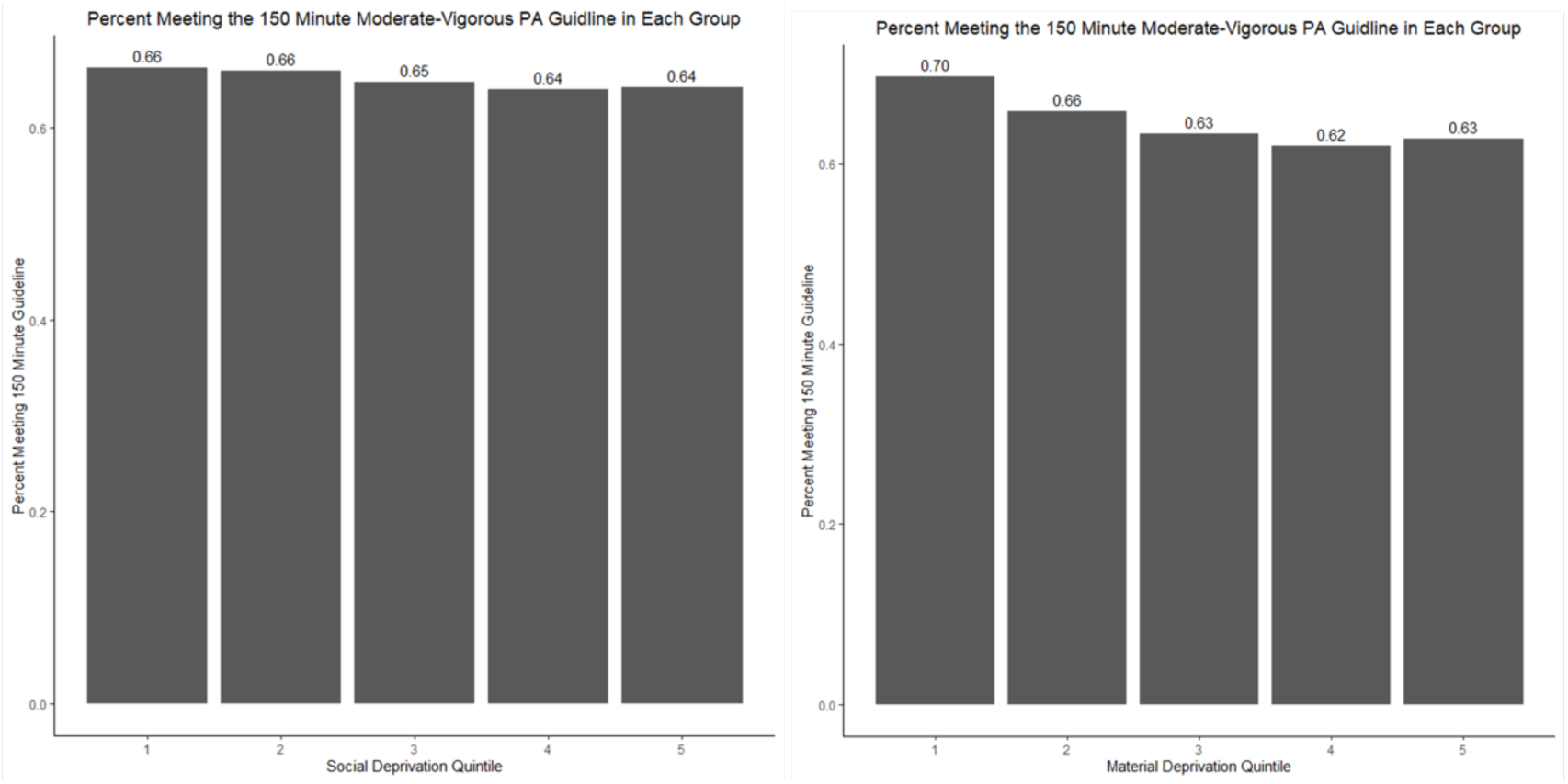
Percent of each quintile reaching 150-minutes of MVPA.

**Figure H3.**
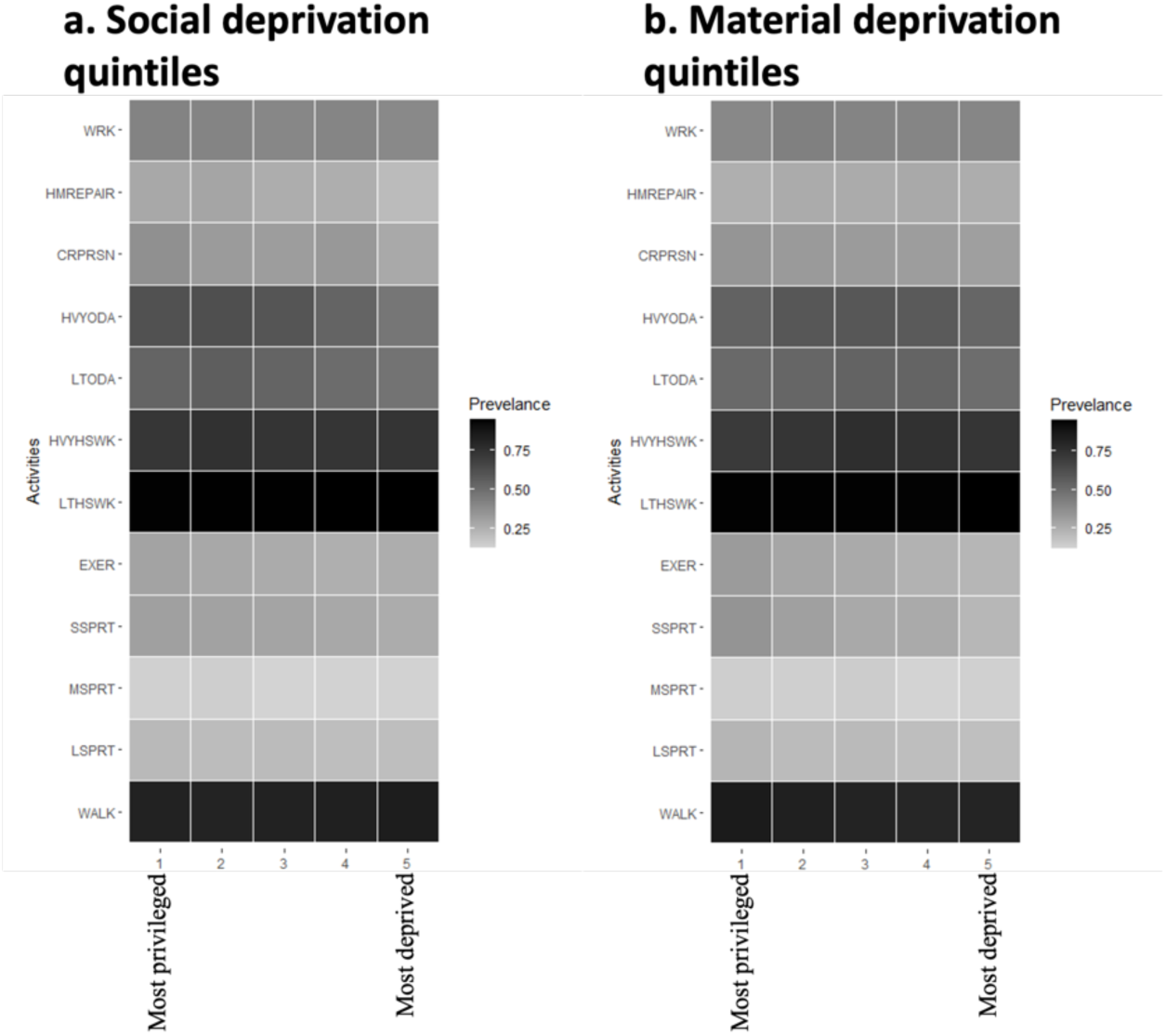
Percentage of each deprivation quintile participating in activities.

## Appendix I. Region and Season Stratification

**Figure I1.**
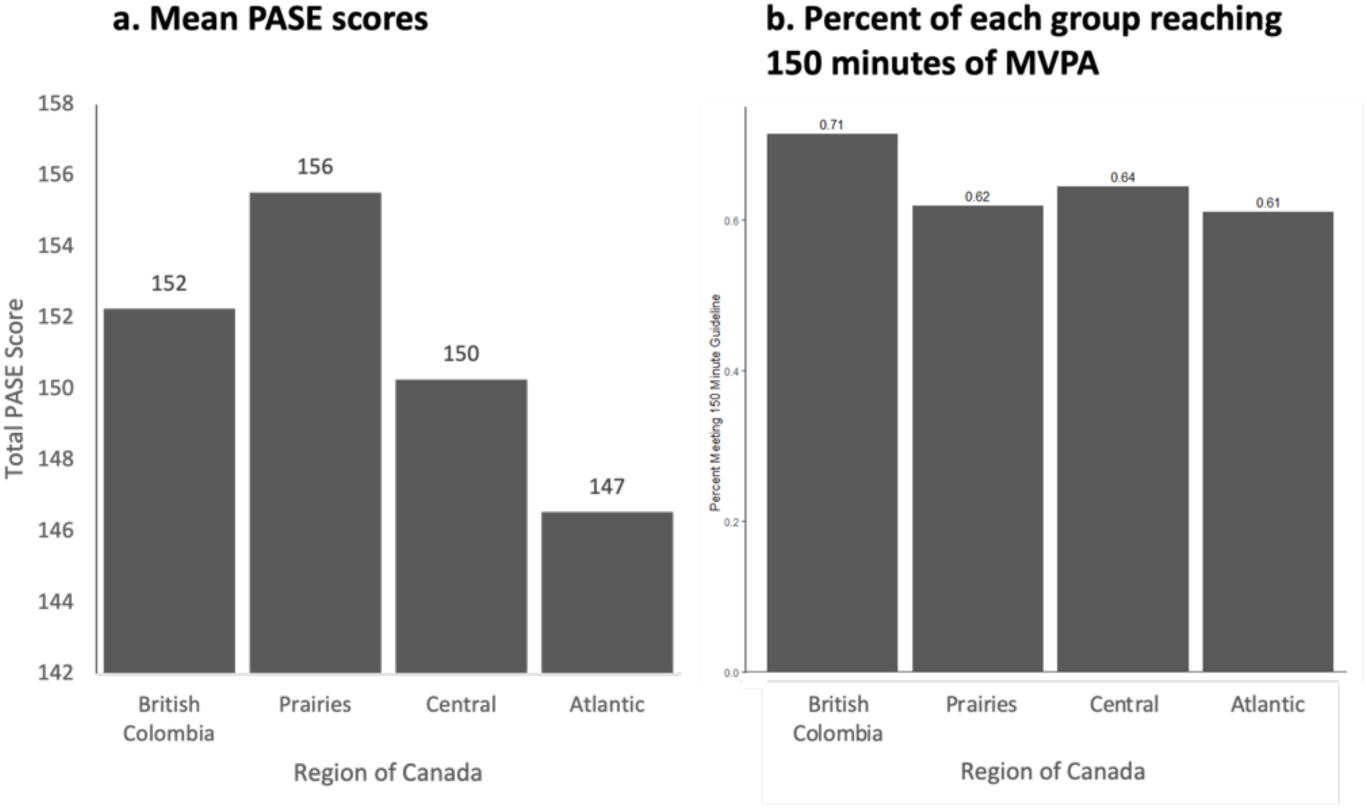
Amount of physical activity by geographical regions in Canada.

**Figure I2.**
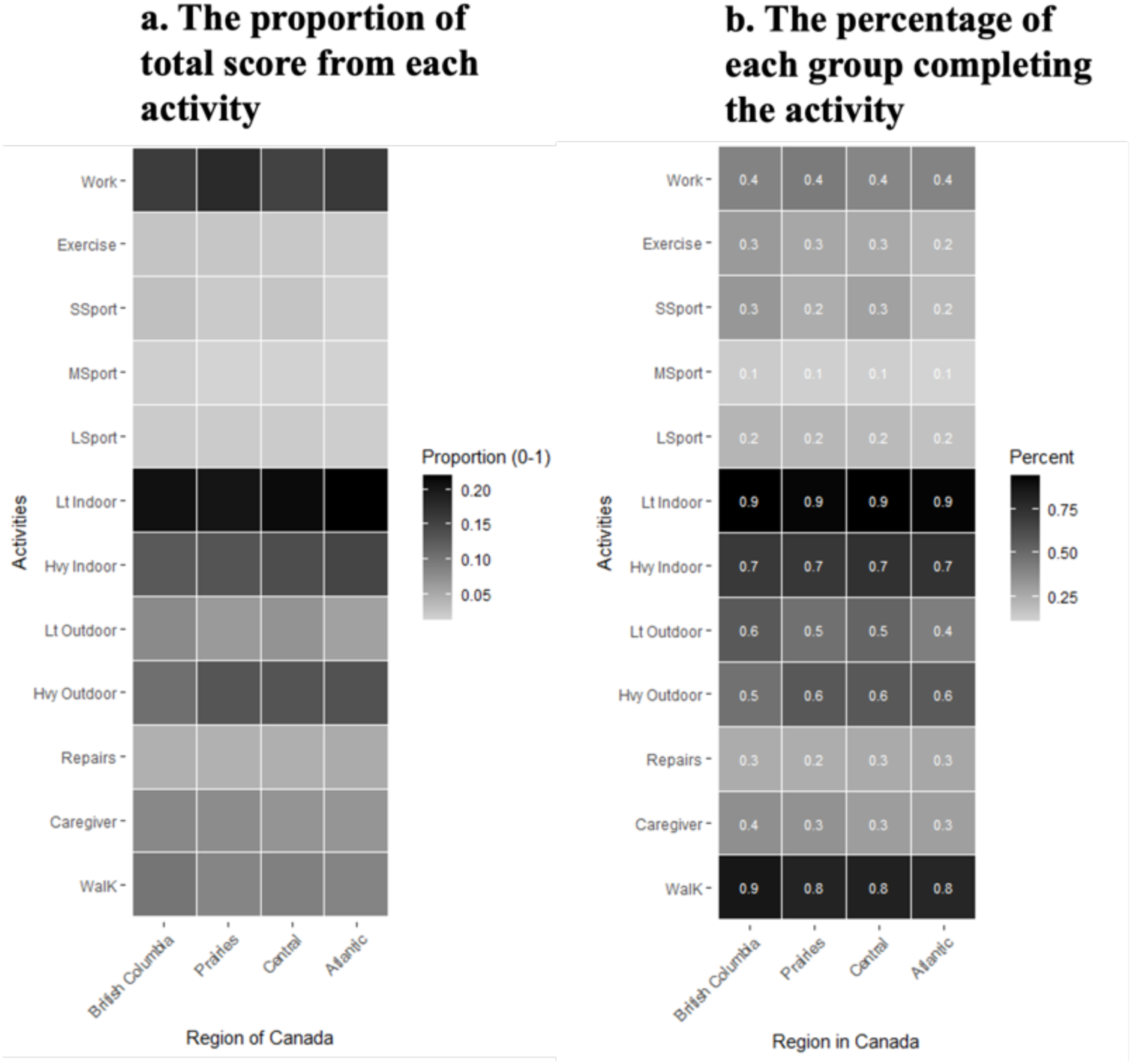
Activity type heat map by region.

**Figure I3.**
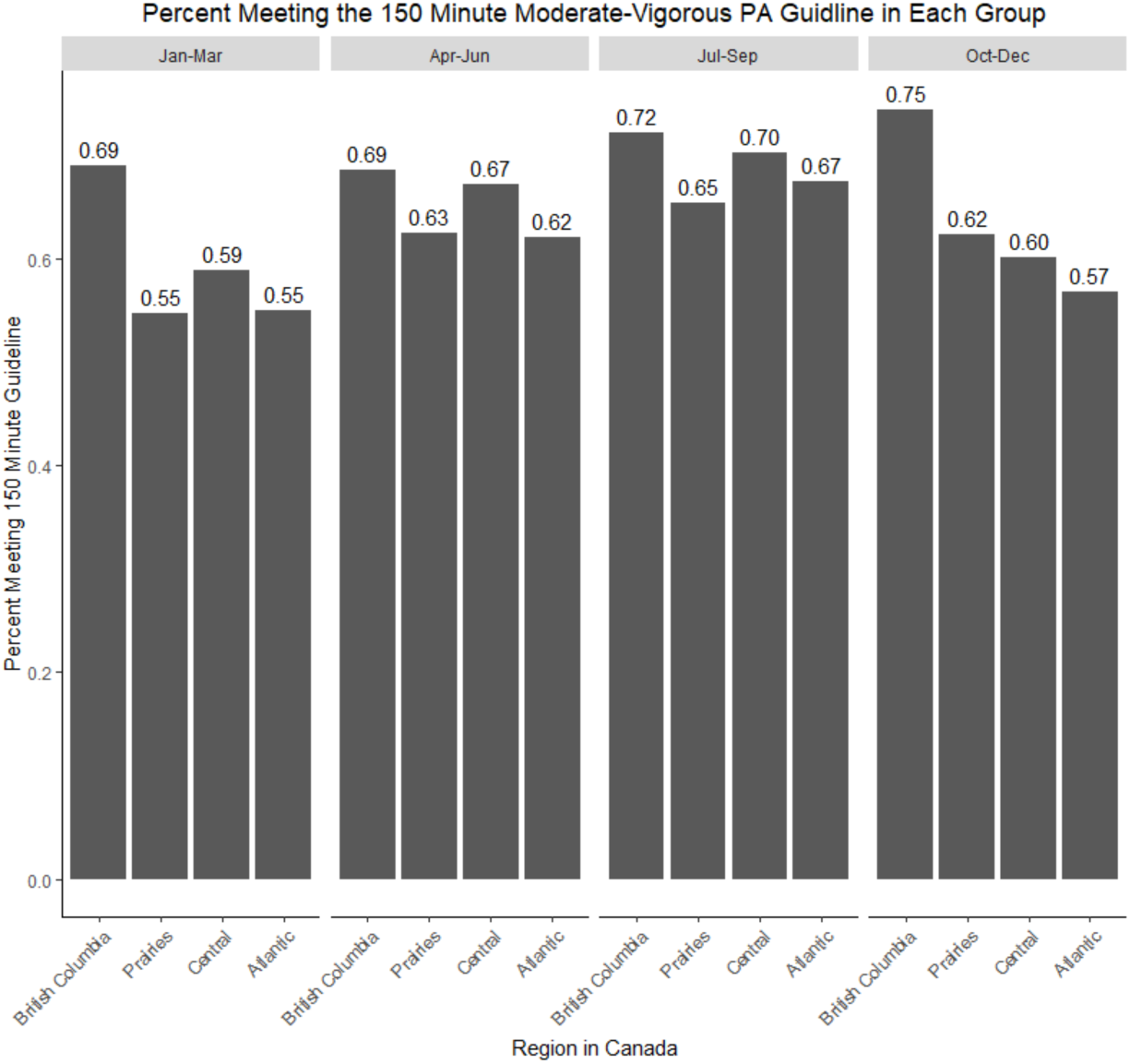
Percentage of each region and season group achieving 150minutes of MVPA.

